# Polygenic Risk and Rare Variants in Endotypes of Idiopathic Pulmonary Fibrosis

**DOI:** 10.1101/2025.05.22.25328177

**Authors:** Anna Duckworth, Leigh Jackson, Harry Green, Atlas Khan, Chen Wang, Andrew Condescu, Gareth Hawkes, Imre Noth, Fernando J Martinez, Ganesh Raghu, Chad A. Newton, Matthew Moll, Michael Cho, Michael Gibbons, Chris J. Scotton, Columbia Genomics Consortium, Krzysztof Kiryluk, Christine Kim Garcia, David Zhang

**Affiliations:** Department of Biomedical and Clinical Sciences, University of Exeter, Exeter, UK; NIHR Exeter Biomedical Research Centre, University of Exeter, Exeter, UK; Academic Department of Respiratory Medicine, Royal Devon University Hospitals NHS Foundation Trust, Exeter, UK; Division of Pulmonary and Critical Care, Department of Medicine, University of Virginia, Charlottesville, VA; Division of Pulmonary, Allergy, and Critical Care, Department of Medicine, University of Massachusetts, Worcester, MA; Division of Pulmonary, Critical Care, and Sleep Medicine, Department of Medicine, University of Washington Medical Center, Seattle, WA; Division of Pulmonary and Critical Care, Department of Medicine, University of Texas Southwestern Medical Center, Dallas, TX; Channing Division of Network Medicine, Brigham and Women’s Hospital, Harvard Medical School, Boston, MA; Division of Pulmonary and Critical Care Medicine, Department of Medicine, Mass General Brigham, Boston, MA; Division of Pulmonary, Allergy, Critical Care, and Sleep Medicine, Department of Veterans Affairs, West Roxbury, MA; Division of Nephrology, Department of Medicine, Columbia University Irving Medical Center, New York, NY; Precision Medicine Institute, Columbia University Irving Medical Center, New York, NY; Division of Pulmonary, Allergy, and Critical Care, Department of Medicine, Columbia University Irving Medical Center, New York, NY

## Abstract

**Background:** Idiopathic pulmonary fibrosis (IPF) and telomere length (TL) are both strongly linked to rare and common genetic variation. Shortened TL itself may be causal for IPF. Whether rare and common variants compete or cooperate to confer genetic risk of IPF uniformly is unknown.

**Methods:** We used whole genome sequencing (WGS) data from a discovery case-control cohort sequenced at Columbia (777 IPF, 2905 controls) and validated findings using WGS data from Trans-Omics for Precision Medicine (TOPMed, 1148 IPF, 5202 controls) and the UK Biobank (UKBB, 2739 IPF, 395331 controls). In all cohorts, we identified rare damaging variants in disease-associated genes and computed control-normalized polygenic risk scores for IPF (IPF-PRS) and telomere length (TL-PRS). Telomere length of blood leukocytes was measured using a qPCR assay for two cohorts. We determined the association of the *MUC5B* rs35705950 polymorphism, an IPF-PRS excluding *MUC5B* (IPF-PRS-no*MUC5B*), and a TL-PRS with IPF risk in the overall cohort and in subgroups stratified by genetic endotypes (rare variant carriers, non-carriers stratified by TL cutoffs). We calculated cross-validated area under the receiver operator curve (AUC) and compared the liability of IPF explained by genetic variables.

**Findings:** We identified independent associations between IPF risk and rare variants, the *MUC5B* SNP, and both polygenic scores in the discovery cohort and replicated these findings in the TOPMed and UKBB cohorts. The adjusted effect size of the TL-PRS, which includes >180 SNPs not previously associated with IPF, was comparable to the IPF-PRS-no*MUC5B* in the discovery (ORTL-PRS 1.63 [95% CI 1.47, 1.81] vs. ORIPF-PRS 1.60 [1.44, 1.77]) and replication cohorts (TOPMed ORTL-PRS 1.47 [1.36, 1.59] vs. ORIPF-PRS 1.37 [1.25, 1.50]; UKBB ORTL-PRS 1.24 [1.19, 1.29] vs. ORIPF-PRS 1.25 [1.21, 1.30]). The TL-PRS incrementally improved disease prediction beyond known IPF common and rare genetic predictors and clinical variables in discovery (combined AUC: 0.89, pDelong = 0.006), TOPMed (combined AUC: 0.89, pDelong = 0.01), and UKBB cohorts (combined AUC: 0.77, pDelong = 0.03). Rare and common variants jointly contributed to genetic liability of IPF. The TL-PRS increased liability of IPF explained by 13% in the discovery cohort and 8% and 13% in the TOPMed and UKBB cohorts, respectively. In IPF subjects with damaging rare variants, the TL-PRS was consistently associated with disease risk whereas the IPF-PRS-no*MUC5B* was not. The TL-PRS also conferred nominally greater odds of disease risk than the IPF-PRS-no*MUC5B* in patients with shorter TL, in the discovery and UKBB cohorts. Together, 23-43% of IPF cases have damaging rare variants or telomeres <10^th^ percentile, where the TL-PRS represents a major unrecognized genetic risk factor.

**Interpretation:** Common and rare genetic variation confer context-specific genetic risk in IPF competitively and cooperatively. In contrast to known IPF common risk variants, the TL-PRS, which includes >180 genetic loci not previously associated with IPF, increases the risk of disease specifically in certain IPF endotypes. Polygenic risk from telomere-associated common variants is a key feature of IPF genetic heterogeneity.

**Funding:** National Institutes of Health (NIH), Medical Research Council (MRC), National Institute for Health and Care Research (NIHR)

**Research in context:** *Evidence before this study:* We performed a search on PubMed for “common variants”, “rare variants”, “polygenic score”, “endotype”, and “IPF” on April 20^th^, 2025, to identify integrated genetic studies of rare and common variants in IPF. We found no original articles that comprehensively examined common and rare IPF variants together. Nearly all articles analyzed common or rare variants in isolation; one article included both common and rare variant analyses but reported results separately without studying their combined effects. We identified numerous reviews that discuss or conceptualize the relative contributions of rare and common genetic risk to IPF risk; no studies provided or cited empiric data. Although “IPF endotypes” have been defined using transcriptomics and proteomics, we did not find any articles that utilized this terminology based on genetic factors, although several studies distinguish IPF with rare variants or telomere length below 10^th^ percentile.

*Added value of this study:* We describe the first comprehensive survey of common and rare risk variants together in IPF and identify subtype-specific genetic risk factors that substantially improve genetic explanation and disease prediction. We uncover complex relationships between polygenic factors and rare variants, including preliminary evidence of polygenic modifiers in IPF carriers of rare damaging variants that might serve as an explanation for incomplete penetrance. Genetic studies in IPF have been limited by sample size leading to “missing heritability”. We demonstrate value in leveraging large-scale genetic studies of causal molecular traits like telomere length to overcome these limitations and improve genetic understanding of IPF. These telomere-associated common variants are context-specific risk factors in certain endotypes, highlighting previously unrecognized genetic heterogeneity that will be important for future discovery of novel, reproducible genetic risk factors in IPF.

*Implications of all the available evidence:* A genetic basis for disease heterogeneity allows for research that focuses on relevant endotypes instead of “all-comer” disease and advances precision medicine approaches to IPF. Polygenic factors that can modify the effects of a high-risk rare variant represent a target for understanding disease-modifying pathways in IPF.

## Introduction

Idiopathic pulmonary fibrosis (IPF) is a lethal, progressive, fibrosing disorder with no known cure. Studies have identified numerous rare and common genetic influences on the risk of developing IPF including those that affect telomere biology^1–6^. Telomeres are hexamer repeats which serve as protective caps at the end of chromosomes and naturally shorten with age or cellular replication^7^. IPF risk variants in telomere maintenance genes, including *TERT, TERC,* and *RTEL1*, cause telomere shortening and have been identified in both genome-wide association studies (GWAS) of common risk variants^8,9^ as well as exome sequencing studies^2,10,11^. Large-scale GWAS of telomere length in healthy individuals have similarly yielded telomere-associated common variants^12,13^. Our previous Mendelian randomization study utilized these telomere-associated SNPs to implicate telomere shortening as being causal for IPF^4^. About 20-30% of IPF patients have short telomere length below the 10^th^ percentile without harboring a rare damaging telomere-gene variant; polygenic risk from telomere-associated SNPs may account for unexplained short telomere length in these cases^6^. Shortened telomere length itself, with or without a rare variant, is linked to worse survival^14–16^, rapid progression^6,17^, adverse response to immunosuppression^18–20^, and complications after transplant^21^. IPF subjects with rare damaging telomere-gene variants or unexplained short telomeres may represent distinct endotypes of disease at higher clinical risk of worse outcomes.

Prior studies have begun to uncover genetic heterogeneity in IPF related to the common *MUC5B* promoter polymorphism^22^, other common risk variants and rare mutations^6,14,23^. Despite these advances, the unaccounted genetic risk in IPF is sizable. Existing studies demonstrate that 75% of cases with familial IPF do not have a genetic diagnosis^6,24^ and that known common risk variants explain only about 10% of disease liability in the general population^25^. However, these studies utilize all-comer IPF cohorts and focus on rare or common variants in isolation, obfuscating the potential role of gene interaction and disease heterogeneity in genetic risk. Comprehensive deconvolution of these factors may help resolve missing heritability in IPF.

In this study, we perform the first integrated assessment of rare damaging variants and common risk variants in IPF by stratifying on heterogenous endotypes of disease. We leverage existing large-scale genetic studies of telomere length heritability to identify novel polygenic risk factors that increase the explained genetic liability of IPF and significantly improve disease prediction. We uncover a complex relationship between common and rare variants in IPF that drives genetic heterogeneity. Our findings support an endotype-aware approach to enhancing genetic discovery in future IPF studies.

## Methods

Please see the online supplement for detailed methods.

### Patient Cohorts

The Columbia discovery cohort with IPF cases and controls has been described previously^6,11^. The institutional review board at Columbia University Medical Center (AAAS0753, AAAS7495, and AAAP0052) approved this study. Replication cohorts with IPF cases and controls from the Trans-Omics for Precision Medicine (TOPMed) program^26^ and the UK Biobank (UKBB)^27^ were used. For the TOPMed cohort, individuals with IPF (phs001607) were classified as cases, and participants of the MESA (phs001416) and Framingham Heart (phs000974) studies were classified as controls. For the UK Biobank cohort, International Classification of Diseases (ICD10) code J84.1 was used to define IPF cases, and exclusion of all J84 codes defined controls.

### Genome Sequencing, Rare Variant Definitions, and Genomic Analysis

Whole genome sequencing (WGS) for the Columbia cohort was performed according to standard protocols on Illumina’s NovaSeq 6000 platform with raw sequencing reads aligned to the hg19 reference genome. WGS data from the discovery cohort and the TOPMed and UK Biobank cohorts were similarly aligned, called and joint genotyped on their respective pipelines. Rare qualifying variants in disease-associated genes predicted to be deleterious were identified using *in silico* tools as previously described^6,11^. Cases and controls within each cohort were pruned for relatedness using *KING*^28^. Principal components were computed using *plink*^29^ to account for population substructure. Genetic ancestry was inferred using *peddy*^30^ using a model trained on 1000 Genomes Project^31^ data.

### Polygenic Risk Scores

We used a pruning and thresholding approach to include autosomal risk variants identified from the summary statistics of GWAS studies that exceeded genome-wide significance to compute polygenic risk scores for IPF and telomere length. We used independent published GWAS studies^9,13^ and identified 16 SNPs related to IPF (**Table S1**) and 190 SNPs related to telomere length (**Table S2**) to compute weighted scores as previously described^6^. Three genome-wide significant loci were associated with both IPF and telomere length **(Figure S1, Table S3).** Given its large effect, we studied the *MUC5B* rs35705950 promoter polymorphism separately from polygenic scores as previously described^32,33^.

For primary analysis, we defined an IPF polygenic risk score using 12 SNPs that excluded the three overlapping telomere-associated SNPs and the *MUC5B* SNP, hereafter termed the “IPF-PRS-no*MUC5B*”. We defined a telomere length polygenic score using 190 SNPs (**Table S2**), hereafter termed the “TL-PRS”. For each cohort, polygenic risk scores (PRS) were normalized to controls and represented as z-scores. For concordance with the IPF-PRS-no*MUC5B*, we negatively inverted the TL-PRS SNP betas so that a higher polygenic score predicts lower telomere length and increased risk of IPF.

For sensitivity analysis we examined multiple alternative polygenic scores **(Table S4)** including an IPF polygenic score that retains three overlapping telomere-associated SNPs and a telomere length polygenic score that excludes these. We also tested a telomere length polygenic score from an independent GWAS study^12^ and an alternative IPF polygenic score using LASSO regression excluding a 500 kb region flanking the *MUC5B* rs35705950 polymorphism^33^. Since both GWAS studies of IPF and telomere length included only European subjects, we also performed sensitivity analyses on European-only and non-European subjects.

### Telomere Length Measurement

For the Columbia cohort and the UK Biobank, peripheral leukocyte telomere length was measured via qPCR as previously described^34,35^. In both cohorts, genomic DNA was isolated from blood leukocytes at time of study enrollment and quality control measures were performed. Age-adjusted percentiles were determined using a multi-ancestry panel of control individuals for the Columbia cohort. For the UK Biobank, internal age-adjusted percentiles were computed using measured ancestry-specific telomere length vigintiles across 5-year age-bands due to differences by race and ethnicity^35^.

### Genetic Liability

To estimate the proportion of disease risk explained by genetic variables, we utilized the liability threshold model which conceptually transforms the observed probability of a binary trait to a continuous liability scale whereby exceeding a critical threshold results in development of that trait^36^. We estimated the genetic liability explained in the general population across a range of disease prevalences^37,38^ for each genetic predictor and each IPF endotype. We estimated the proportion of variance explained by each genetic predictor by calculating the correlation attributed to the predictor in a linear model and comparing full and reduced models adjusted for other genetic predictors, age, sex, and principal components of ancestry. We converted the proportion of variance explained to a liability scale accounting for ascertainment in the case-control cohort across a range of observed prevalences of IPF in the general population as previously described^39^.

### Statistics

We performed logistic regression to assess the association of genetic variables with IPF diagnosis in each cohort adjusting for age, sex, and principal components of ancestry. We selected five principal components based on visualization of flattening of variance explained on scree plot **(Figure S2)**. We assessed gene interaction by assessing the significance of pair-wise interaction terms between genetic variables. To assess disease prediction, we calculated area under the receiver operating curve (AUC) with 10-fold cross-validation using R packages *pROC* and *caret*. Comparison of AUCs was performed using DeLong’s test.

Comparisons of non-parametric continuous data were performed using Wilcoxon rank sum test for two groups or Kruskal-Wallis test for multiple groups. For meta-analyses we used a random-effects model using R package *metafor*.

To estimate genetic correlation between traits, we performed cross-trait linkage disequilibrium (LD) score regression^40^ using published GWAS summary statistics. To minimize bias from population stratification, we focused on studies of individuals of European ancestry. We used LD scores estimated from the 1000 Genomes Project^31^ European reference panel for regression weights.

All p-values less than 0.05 were considered significant with Bonferroni correction applied for multiple comparisons. Statistical analyses were performed using R statistical analysis software, version 4.4.0 (www.r-project.org).

## Results

### Subject characteristics

The discovery cohort included 777 unrelated IPF cases and 2905 non-IPF controls as previously described^11^. Case-control cohorts from TOPMed and the UK Biobank (UKBB) were used for replication. The TOPMed cohort included 1148 unrelated IPF cases and 5202 unrelated, population-based controls from the MESA and Framingham Heart studies as previously described^11^. The UK Biobank cohort included 2739 unrelated IPF cases and 395,331 unrelated controls **(Figure 1A)**. The IPF cohorts were older (median age 64-67), male-predominant (61-72%), and mostly of European ancestry (84-96%) **(Table S6)**.

**Figure 1.**
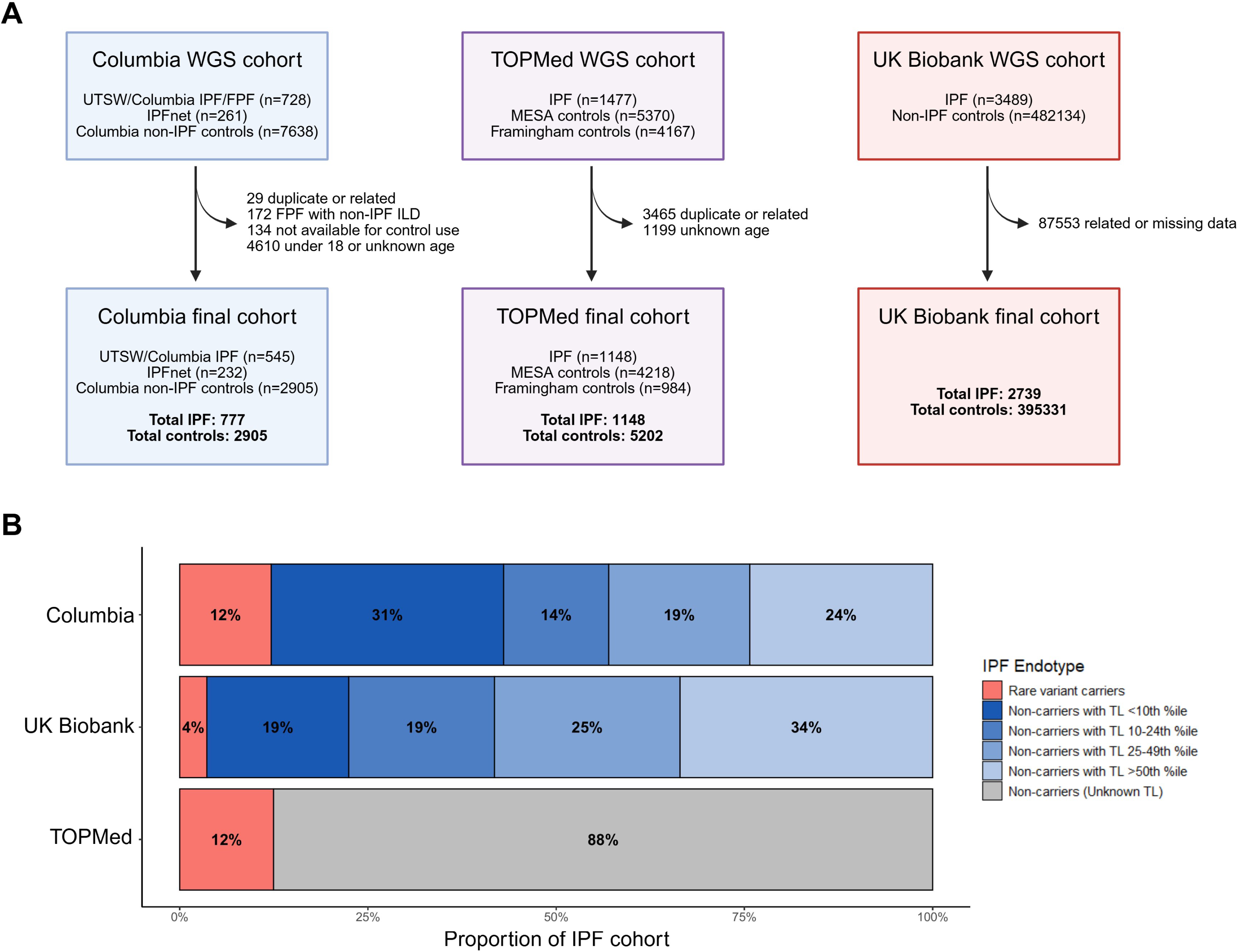
Study cohorts and proportion of IPF endotypes. **A.** Study flow diagram of IPF cohorts with schematic accounting of cases and controls. **B.** Relative proportion of each IPF cohort classified by endotype. Rare variant carriers include patients with inherited rare damaging mutations in IPF-associated genes including *TERT, TERC, PARN, RTEL1, DKC1, TINF2, NAF1, SFTPC, SFTPA1/2,* and *KIF15*. Peripheral blood telomere length measured by qPCR and presented as age-adjusted percentiles. Individuals without rare variants were subdivided based on telomere length: <10^th^ percentile, 10-24^th^ percentile, 25-49^th^ percentile, and >50^th^ percentile. Telomere length percentiles are not available for the TOPMed cohort. Abbreviations: WGS, whole genome sequencing; UTSW, University of Texas Southwestern; FPF, familial pulmonary fibrosis; IPFnet, IPF Clinical Research Network; MESA, Multi-Ethnic Study of Atherosclerosis; TL, telomere length.

### Rare variants, telomere length, and endotype classification

We identified carriers of rare deleterious qualifying variants in IPF-associated genes (*TERT, TERC, RTEL1, PARN, DKC1, TINF2, NAF1, ZCCHC8, SFTPC, SFPTA1/2*, and *KIF15*) as previously described^6^ in all cohorts. In total we identified 94 (12%) carriers in the Columbia cohort, 143 (12%) carriers in the TOPMed cohort, and 108 (3.9%) in the UKBB cohort (**Table S7**). Telomere length measurement was available for the Columbia and UKBB cohorts. For these cohorts, we subdivided IPF cases into endotypes including rare variant carriers, and non-carriers stratified by TL (<10^th^ percentile, 10-24^th^ percentile, 25-49^th^ percentile, and >50^th^ percentile) (**Figure 1B, Table S8**). Most IPF non-carriers had TL below average for age; rare variant carriers and non-carriers with TL <10^th^ percentile represented 23-43% of the IPF cases.

### Association of polygenic scores with IPF

We found a significant difference in the distribution of all PRS between IPF and controls (all p<0.0001) in the Columbia, TOPMed, and UKBB cohorts (**Figure S3**). There was heterogeneity in the TL-PRS amongst IPF cases with different telomere lengths (Kruskal-Wallis p = 5.9×10^−5^, **Figure S6**). In multivariable analysis, we find significant independent associations between IPF risk and rare variants (OR 12.0, 95% CI 7.57-19.5, p=1.0×10^−24^), *MUC5B* SNP (OR 2.00, 95% CI 1.85-2.15, p=4.0×10^−72^), the IPF-PRS-no*MUC5B* (OR 1.60, 95% CI 1.44-1.77, p=9.4×10^−19^), and the TL-PRS (OR 1.63, 95% CI 1.47-1.81, p=5.5×10^−21^) adjusting for age, sex, genetic variables, and principal components of ancestry (**Table 1**). We did not find consistent evidence of a significant pair-wise interaction term between genetic variables (**Figure S7**). In the discovery cohort, AUC analysis demonstrated improved disease prediction when genetic variables were added to a clinical model of age and sex (**Figure 2A-B**). The inclusion of the TL-PRS in a combined model with clinical variables and common and rare variables significantly improved disease prediction in the discovery cohort (AUC: 0.88 to 0.89, DeLong’s p = 0.006). Analysis in TOPMed and UKBB replication cohorts affirmed that inclusion of clinical variables with all rare and common genetic variables, including the TL-PRS, led to the best prediction of IPF (TOPMed combined AUC = 0.89, UKBB combined AUC = 0.77) (**Table 1**, **Figure 2C-F**).

**Figure 2.**
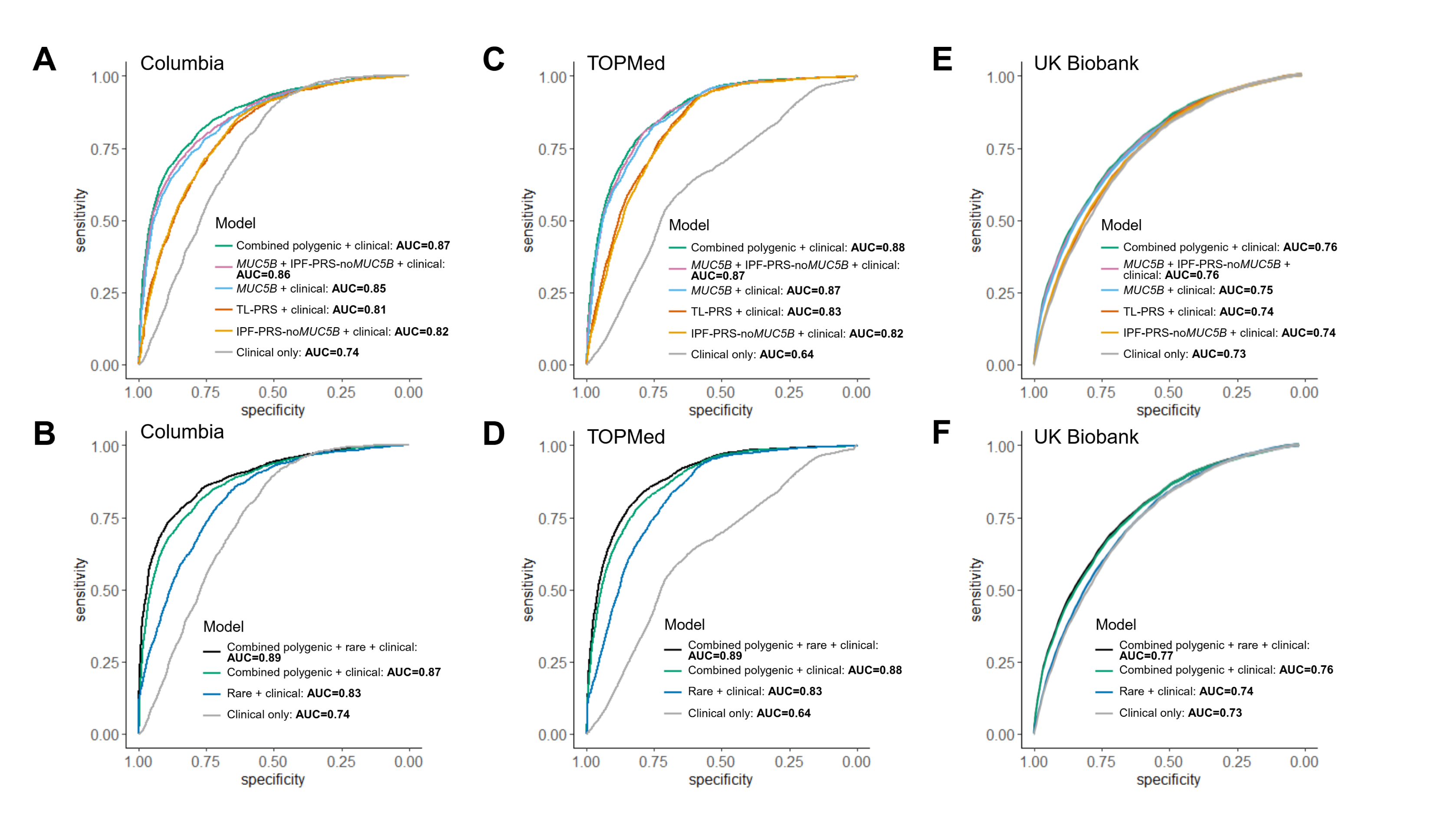
Receiver operator curves (ROC) of clinical and genetic predictors of IPF. Data shown using **(A, B)** Columbia cohort, **(C, D)** TOPMed cohort, and **(E, F)** the UK Biobank. Area under ROC calculated with 10-fold cross validation. All models with genetic predictors include 5 PC of ancestry. Clinical model includes age and sex. **(A, C, E)** ROC curves combining common variant polygenic risk scores and clinical predictors. In all cohorts, DeLong’s test demonstrates significant improvement in prediction with the addition of individual genetic predictors to clinical predictors (all p < 2.0 x 10^−16^). **(B, D, F)** A model combining common and rare genetic variables with clinical predictors results in the best disease prediction (Columbia AUC 0.89, TOPMed AUC 0.89, UKBB AUC 0.77). The TL-PRS incrementally improves disease prediction beyond known IPF common and rare variants and clinical predictors (Columbia pDelong = 0.006; TOPMed pDelong = 0.01, UKBB pDelong = 0.03). *MUC5B* SNP, rs35705950 polymorphism; IPF-PRS-no*MUC5B,* IPF polygenic risk score excluding *MUC5B* SNP and overlapping telomere-associated loci; TL-PRS, telomere length polygenic score.

**Table 1.** Associations between polygenic scores and IPF in multiple cohorts.

| Variable | Columbia |  |  | TOPMed |  |  | UK Biobank |  |  |
| --- | --- | --- | --- | --- | --- | --- | --- | --- | --- |
|  | OR (95% CI) | P | AUC | OR (95% CI) | P | AUC | OR (95% CI) | P | AUC |
| Clinical model | - | - | 0.74 | - | - | 0.64 | - | - | 0.73 |
| Age | 1.06 (1.05, 1.07) | 6.1x10<sup>-71</sup> |  | 1.02 (1.01, 1.03) | 5.9x10<sup>-10</sup> |  | 1.12 (1.11, 1.12) | 1.3x10<sup>-274</sup> |  |
| Male sex | 1.85 (1.54, 2.22) | 9.9x10<sup>-12</sup> |  | 2.43 (2.12, 2.79) | 7.0x10<sup>-37</sup> |  | 1.75 (1.62, 1.89) | 1.9x10<sup>-45</sup> |  |
| *MUC5B* rs35705950 | 2.08 (1.95, 2.23) | 5.5x10<sup>-104</sup> | 0.77 | 1.87 (1.78, 1.97) | 6.1x10<sup>-123</sup> | 0.85 | 1.44 (1.40, 1.48) | 4.1x10<sup>-136</sup> | 0.61 |
| IPF-PRS-no*MUC5B* | 1.61 (1.49, 1.76) | 2.4x10<sup>-29</sup> | 0.71 | 1.45 (1.35, 1.57) | 2.4x10<sup>-21</sup> | 0.80 | 1.25 (1.20, 1.29) | 1.8x10<sup>-33</sup> | 0.57 |
| TL-PRS | 1.55 (1.43, 1.68) | 4.0x10<sup>-26</sup> | 0.71 | 1.52 (1.42, 1.63) | 1.4x10<sup>-33</sup> | 0.81 | 1.24 (1.20, 1.29) | 9.2x10<sup>-30</sup> | 0.57 |
| Rare variants | 8.78 (6.01, 13.0) | 2.6x10<sup>-28</sup> | 0.69 | 13.8 (9.49, 20.6) | 1.6x10<sup>-40</sup> | 0.79 | 3.11 (2.52, 3.78) | 7.8x10<sup>-28</sup> | 0.54 |
| *MUC5B* rs35705950 + clinical | 2.02 (1.88, 2.17) | 1.3x10<sup>-85</sup> | 0.85 | 1.86 (1.77, 1.97) | 2.3x10<sup>-116</sup> | 0.87 | 1.45 (1.41, 1.49) | 4.2x10<sup>-140</sup> | 0.75 |
| IPF-PRS-no*MUC5B* + clinical | 1.63 (1.49, 1.79) | 3.0x10<sup>-26</sup> | 0.82 | 1.43 (1.33, 1.55) | 3.6x10<sup>-19</sup> | 0.82 | 1.25 (1.21, 1.30) | 1.2x10<sup>-33</sup> | 0.74 |
| TL-PRS + clinical | 1.58 (1.45, 1.73) | 2.4x10<sup>-24</sup> | 0.81 | 1.52 (1.42, 1.63) | 1.8x10<sup>-32</sup> | 0.83 | 1.24 (1.19, 1.28) | 3.6x10<sup>-29</sup> | 0.74 |
| Rare variants + clinical | 12.1 (7.88, 18.8) | 2.8x10<sup>-29</sup> | 0.83 | 15.5 (10.6, 23.4) | 6.0x10<sup>-42</sup> | 0.83 | 3.23 (2.62, 3.95) | 2.6x10<sup>-29</sup> | 0.74 |
| Combined polygenic + clinical* |  |  | 0.87 |  |  | 0.88 |  |  | 0.76 |
| *MUC5B* rs35705950 | 2.01 (1.87, 2.16) | 6.2x10<sup>-77</sup> |  | 1.84 (1.74, 1.94) | 3.3x10<sup>-107</sup> |  | 1.45 (1.41, 1.49) | 4.7x10<sup>-140</sup> |  |
| IPF-PRS-no*MUC5B* | 1.56 (1.41, 1.72) | 4.3x10<sup>-18</sup> |  | 1.36 (1.24, 1.48) | 5.8x10<sup>-12</sup> |  | 1.25 (1.20, 1.29) | 2.2x10<sup>-33</sup> |  |
| TL-PRS | 1.64 (1.49, 1.82) | 1.6x10<sup>-22</sup> |  | 1.46 (1.35, 1.57) | 4.0x10<sup>-22</sup> |  | 1.24 (1.19, 1.29) | 4.1x10<sup>-29</sup> |  |
| Combined polygenic + rare + clinical* |  |  | 0.89 |  |  | 0.89 |  |  | 0.77 |
| *MUC5B* rs35705950 | 2.00 (1.85, 2.15) | 4.0x10<sup>-72</sup> |  | 1.86 (1.77, 1.97) | 7.8x10<sup>-104</sup> |  | 1.45 (1.41, 1.49) | 4.4x10<sup>-140</sup> |  |
| IPF-PRS-no*MUC5B* | 1.60 (1.44, 1.77) | 9.4x10<sup>-19</sup> |  | 1.37 (1.25, 1.50) | 7.6x10<sup>-12</sup> |  | 1.25 (1.21, 1.30) | 1.0x10<sup>-33</sup> |  |
| TL-PRS | 1.63 (1.47, 1.81) | 5.5x10<sup>-21</sup> |  | 1.47 (1.36, 1.59) | 1.8x10<sup>-21</sup> |  | 1.24 (1.19, 1.29) | 5.0x10<sup>-29</sup> |  |
| Rare variants | 12.0 (7.54, 19.5) | 1.0x10<sup>-24</sup> |  | 18.2 (12.0, 28.2) | 2.5x10<sup>-40</sup> |  | 3.28 (2.66, 4.01) | 9.7x10<sup>-30</sup> |  |

All associations with genetic predictors are adjusted for 5 PCs of ancestry. Common variant genetic variables are coded as control-normalized z-transformed scores; each one unit increase in OR represents one standard deviation increase in genetic predictor. Rare variant variable is coded as presence or absence; each one unit increase in OR represents presence of rare variant. \*Model includes genetic predictors, age, sex, and 5 principal components of ancestry. IPF-PRS-no*MUC5B*, IPF polygenic risk score excluding the *MUC5B* rs35705950 polymorphism and overlapping telomere-associated loci (see supplementary Methods); TL-PRS, telomere length polygenic score.

### Endotype-specific polygenic effects on IPF risk

Since we found independent effects from polygenic scores, we explored their relevance to different IPF endotypes, including rare variants carriers and non-carriers with different telomere lengths. In the discovery cohort (**Figure 3A**), the *MUC5B* polymorphism was similarly associated with IPF across all endotypes (overall OR*MUC5B* 2.02, 95% CI [1.88, 2.17]). The IPF-PRS-no*MUC5B* was associated with IPF risk amongst non-carriers (ORIPF-PRS 1.69, 95% CI [1.54, 1.86]) but *not* amongst rare variant carriers (ORIPF-PRS 1.23, 95% CI 1.00, 1.51]). The TL-PRS was associated with IPF risk in rare variant carriers and non-carriers, with the highest odds ratio in the TL <10^th^ percentile group (ORTL-PRS 2.02, 95% CI [1.76, 2.33]) and the lowest in the TL >50^th^ percentile group (ORTL-PRS 1.23, 95% CI [1.06, 1.44]). In sensitivity analyses, the use of alternative polygenic scores, including an IPF-PRS that retains the overlapping telomere-associated loci, yielded similar endotype-specific associations (**Figure S8**).

**Figure 3.**
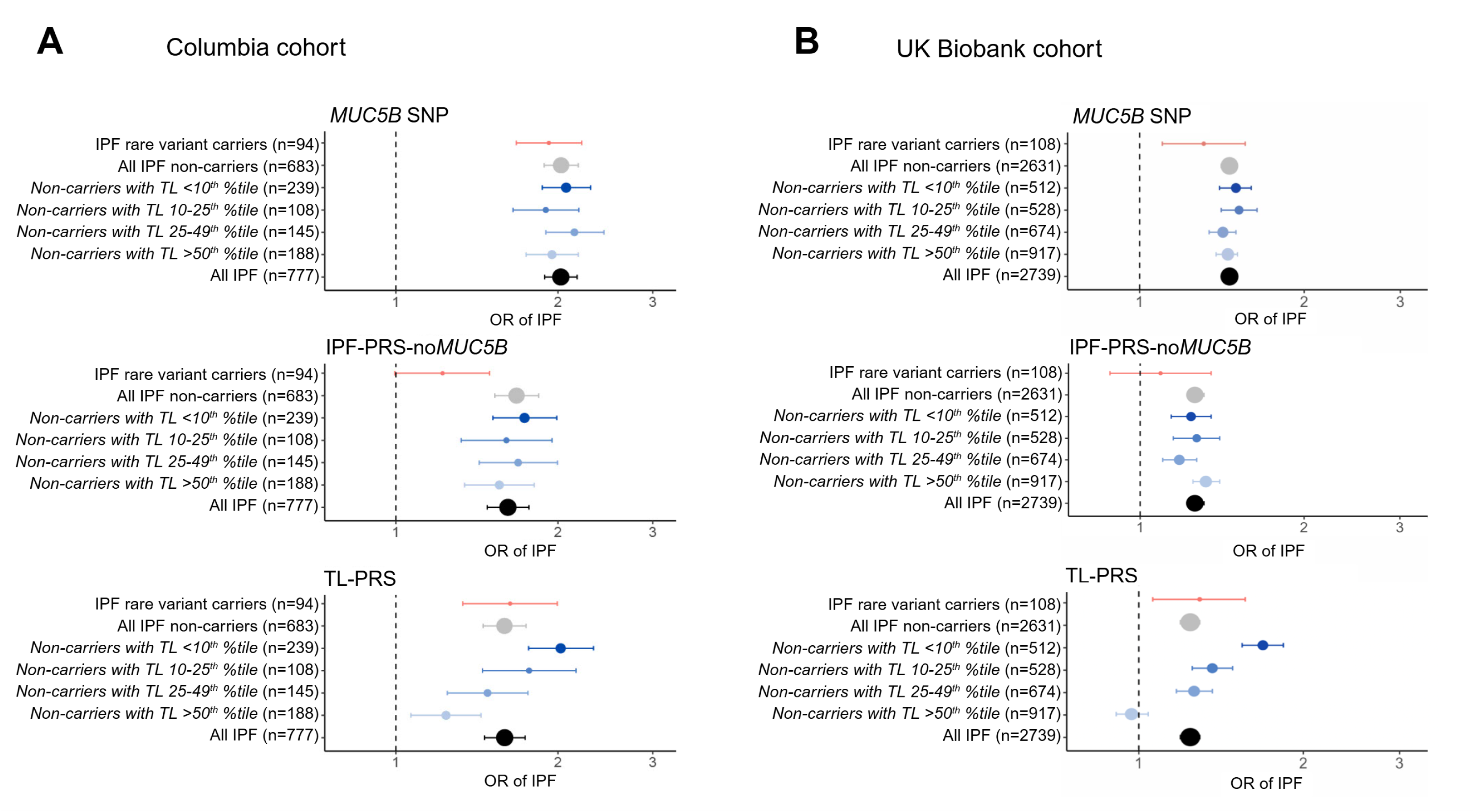
Associations of genetic predictors and polygenic scores with IPF risk across endotypes of disease. Odds ratios and 95% confidence intervals shown using data from **(A)** Columbia cohort on left and **(B)** UK Biobank on right. All associations adjusted for age, sex and 5 PCs of ancestry. Genetic predictors are displayed as control-normalized z-transformed scores; each one unit increase in OR represents one standard deviation increase in genetic predictor. The top panels show a similar effect of the *MUC5B* polymorphism on IPF risk across IPF endotypes. The middle panels show a diminished effect of the IPF-PRS-no*MUC5B* on disease risk in rare variant carriers. The bottom panels show heterogenous effects of the TL-PRS on IPF risk in rare variant carriers and non-carriers with various telomere lengths. For both the Columbia cohort and the UK Biobank, the TL-PRS had the greatest effect in non-carriers with telomere lengths <10^th^ percentile.

Replication analysis in the TOPMed and UKBB cohorts confirmed endotype-specific genetic risk factors. The effect of IPF-PRS-no*MUC5B* on disease risk was similarly attenuated in rare variant carriers vs non-carriers in the TOPMed replication cohort (carriers ORIPF-PRS 1.30, 95% CI [1.08, 1.57]) (**Figure S9**), and in the UKBB replication cohort (carriers ORIPF-PRS 1.07, 95% CI [0.88, 1.30]) (**Figure 3B**). The TL-PRS had a similarly heterogenous effect on disease risk in non-carriers in the UKBB with significant associations in those with shortest TL <10^th^ percentile (ORTL-PRS 1.70, 95% CI [1.56, 1.85]), but not those with TL >50^th^ percentile (ORTL-PRS 0.95, 95% CI [0.93, 1.06]). Overall, the TL-PRS was consistently and independently associated with IPF risk in rare variant carriers and non-carriers with TL <10^th^, between 10-24^th^, and between 25-49^th^ percentiles in discovery and UKBB cohorts, representing 65-75% of cases.

To understand the portability of these polygenic scores across ancestry, we performed a stratified analysis based on European ancestry. In European individuals, all associations remained consistent **(Figure S10).** In non-European individuals, the *MUC5B* SNP associations remained consistent across cohorts in IPF overall, but the IPF-PRS-no*MUC5B* and TL-PRS associations were not consistent although the precision of estimates was limited by small sample size (**Figure S11**).

### Proportion of liability explained by common and rare variants

To conceptualize the relative contributions from each genetic variable, we compared the proportion of IPF risk conferred using liability estimates^36^ across a range of reported IPF prevalences^37^ in the general population as well as in individuals older than 65^38^ **(Figure 4A)**.

**Figure 4.**
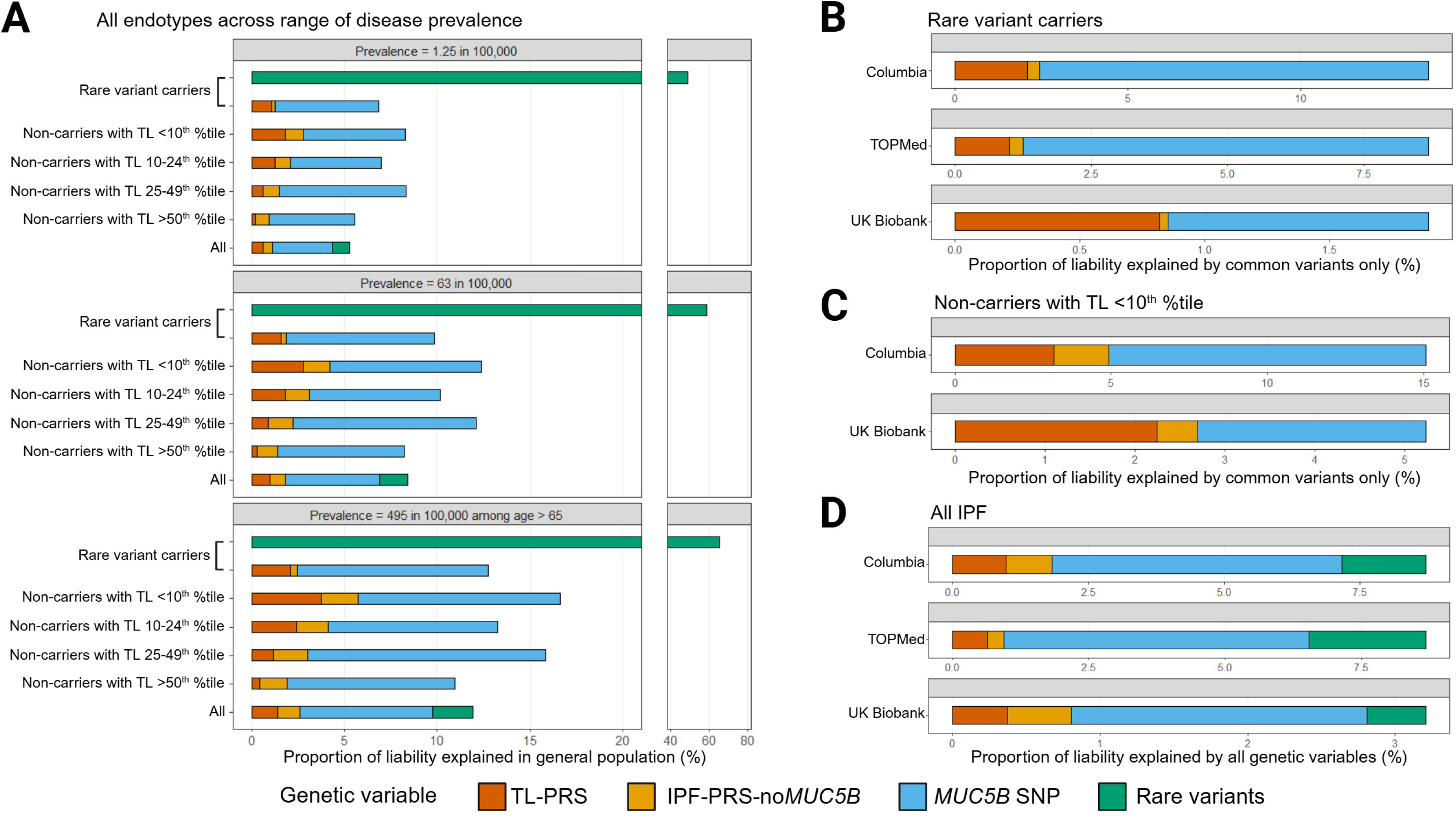
Liability of developing IPF in the general population explained by known rare and common genetic risk factors. Stacked bar plots demonstrate estimated genetic liability explained by rare and common variant risk factors. **(A)** Using the Columbia cohort, proportion of liability estimated by each genetic predictor is shown assuming a range of observed prevalences of IPF. There is a dominant effect of rare variants on liability explained in IPF rare variant carriers. The *MUC5B* SNP explains the most liability amongst IPF non-carriers. The TL-PRS accounts for more liability in non-carriers with shorter telomere lengths than those with longer telomere length. Data from the TOPMed and UK Biobank cohorts **(B, C, D)** are compared to the Columbia cohort assuming a disease prevalence of 63 in 100,000. Panel **B** shows the liability explained by common variants in rare variant carriers; in all cohorts the TL-PRS explains a greater proportion of genetic liability than the IPF-PRS-no*MUC5B*. Panel **C** shows the liability explained by common variants in IPF non-carriers with telomere length <10^th^ percentile; in both cohorts the TL-PRS explains more liability than the IPF-PRS-no*MUC5B*. Panel **D** shows liability explained by all genetic variables in IPF overall. Across multiple cohorts, rare variants and the TL-PRS together improve the genetic liability explained by ∼30%.

For IPF rare variant carriers, we reported the liability explained by rare variants and polygenic factors separately due to potential collinearity between genetic factors in this group. Rare variants alone explained most of the liability (40-60%) far exceeding polygenic factors in rare variant carriers. When comparing polygenic risk factors in rare variant carriers, the TL-PRS explained more disease liability than the IPF-PRS-no*MUC5B* (1.1-2.0% vs 0.2-0.4%).

Amongst IPF subjects without a rare damaging variant, the *MUC5B* SNP explained a major fraction of liability. The TL-PRS explained more liability than the IPF-PRS-no*MUC5B*, especially for non-carriers with TL <10^th^ percentile (1.8-3.6% vs 1.0-2.0%), and those with TL 10-24^th^ percentile (1.2-2.3% vs 0.9-1.8%). The endotype-specific genetic liability explained by TL-PRS persisted in sensitivity analyses using alternative polygenic scores and in non-Europeans **(Figures S13, S14, and S15)**.

Across all IPF cases, we identified the overall liability explained by *MUC5B* polymorphism (3.1-6.9%), rare variants (0.9-2.1%), the IPF-PRS-no*MUC5B* (0.5-1.2%), and the TL-PRS (0.6-1.3%). The TL-PRS, comprising >180 SNPs not previously associated with IPF, increased the genetic liability of IPF explained overall by 13%. We also observed similar overall and endotype-specific trends in genetic liability explained in the TOPMed and UKBB replication cohorts **(Figure 4B-D).**

### Genetic correlation

To assess the genetic overlap between IPF and telomere length in the context of other epidemiologically-associated traits, we estimated pair-wise genetic correlation amongst IPF^9^, telomere length^13^, smoking^41^, GERD^42^, monocyte count^43^, rheumatoid arthritis^44^, and forced vital capacity (FVC)^45^ utilizing summary statistics from large-scale GWAS studies for all traits (**Figure S16, S17**). Telomere length was the only trait with significant genetic correlation with IPF after correcting for multiple comparison testing (r=-0.35, p=3.6×10^−6^).

## Discussion

Our study utilized three independent cohorts comprising over 4,500 IPF cases and 400,000 controls to systematically characterize the interplay of rare and common genetic risk factors in genetic endotypes of IPF **(Figure 5)**. We observed outsized effects from polygenic risk of telomere shortening, especially in 23-43% of IPF cases with a damaging rare variant or very short telomere length. Our study highlights the utility of understanding genetic heterogeneity for uncovering novel IPF genetic risk factors.

**Figure 5.**
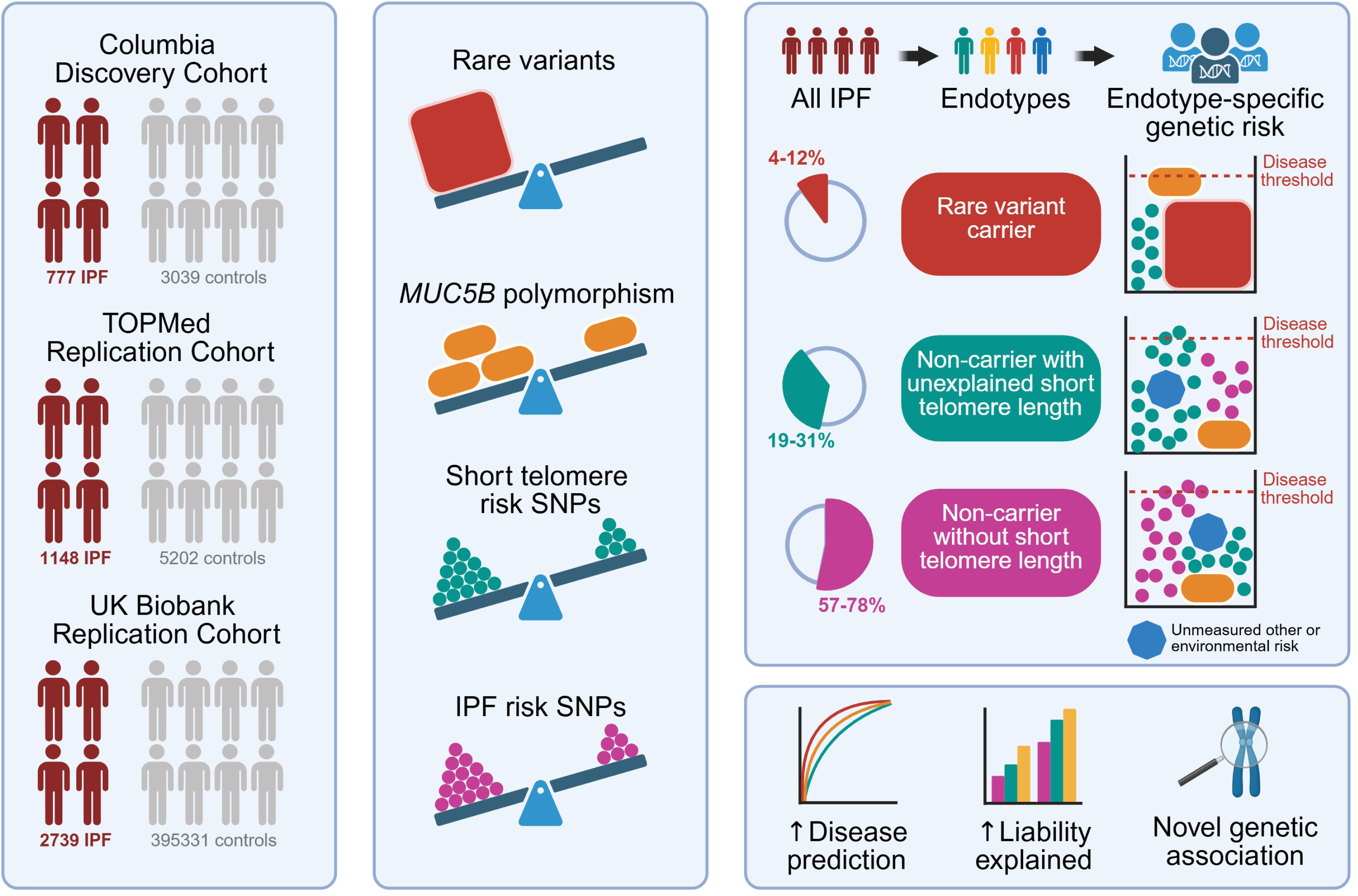
Graphical summary of study. Genetic association of rare and common variants stratified by genetic endotype performed across three independent case-control cohorts of IPF comprising >4,500 IPF cases and >400,000 controls. The *MUC5B* polymorphism (*yellow oval*), short telomere risk SNPs (*green circle*), IPF risk SNPs (*purple circle*), and rare variants (*red box*) were all independently associated with risk of IPF in all cohorts. Three genetic endotypes of IPF were identified including rare variant carriers (4-12%), non-carriers with unexplained short telomere length (19-31%), and other non-carriers (57-78%). Endotype-specific combinations of genetic risk factors were identified. Short telomere risk SNPs (*green circle*) include >180 SNPs not previously associated with IPF and contribute greater proportion of genetic risk than IPF risk SNPs (*purple circle*) in IPF with unexplained short telomeres or rare variants. Accounting for all common and rare genetic risk factors improved disease prediction in all cohorts and explained more liability of disease.

Studying the genetic basis of associated molecular phenotypes can substantially improve mechanistic understanding of diseases and lead to therapies that focus on treating causal traits. Successful examples of this approach include genetic studies of low-density lipoprotein (LDL) cholesterol levels for coronary artery disease^46^, fetal hemoglobin levels for hemoglobinopathies^47^, or serum IgA levels for IgA nephropathy^48^.

Similarly, our approach leveraged telomere length GWAS studies involving hundreds of thousands of participants to identify novel IPF genetic associations, capitalizing on robust polygenic scores from well-powered analyses. Genetic studies of molecular traits like telomere length often have larger sample sizes and yield more informative genetic findings compared to studies of a rare diagnosis like IPF. Alternative polygenic scores using thousands of SNPs for IPF risk^33^ and 20 SNPs^12^ for telomere length heritability affirmed the value of a well-powered polygenic score and supported our primary conclusions.

We highlight a complex relationship between polygenic risk factors and rare variants in IPF. Non-*MUC5B* IPF common risk variants have diminished effect in carriers of rare damaging variants, suggesting that these two genetic variables “compete” to confer risk of IPF. In contrast, the *MUC5B* SNP and telomere-associated common variants “cooperate” with rare risk variants. Most of these rare variants are also found in telomere maintenance genes, suggesting that rare and common variants that govern telomere length confer IPF risk additively. Rare telomere-gene variants confer upwards of 40-fold increased odds of IPF and are linked to monogenic forms of disease^10,11^ but feature variable penetrance. Studies of familial pulmonary fibrosis kindreds often identify rare variant carriers who are asymptomatic^24^ and have found that the presence of the *MUC5B* SNP is linked to radiographic evidence of early fibrotic lung disease^49^. Polygenic modification has also been proposed to explain incomplete penetrance in other monogenic disorders including cardiovascular disease^50^, hereditary cancer^50^, metabolic disease^51^, and chronic kidney disease^52^. Our findings pave the way to study polygenic modifiers as an explanation for variable penetrance in families of IPF rare variant carriers.

Historically, the genetic link between telomere shortening and IPF has been attributed to rare variants^1–3,5,53^. We find that telomere-associated common variants can explain a substantial portion of IPF genetic risk as well. In IPF patients with unexplained short telomere length, the polygenic risks of IPF and of telomere shortening converge, highlighting a parallel genetic pathway for future studies of disease mechanism of this specific endotype. Furthermore, telomere length testing and genetic testing for IPF are both available clinically^54^, allowing for potential precision medicine based clinical care in the future. Interestingly, telomere-associated common variants confer risk of IPF even amongst those with telomere lengths above the 10^th^ percentile, up to the 50^th^ percentile. A spectrum of telomere-related polygenic risk appears to be relevant in the 65-75% of IPF patients with telomere lengths shorter than expected for a given age. These genetic influences on telomere length are likely independent of epigenetic modification of telomere “set-length” itself^55^ by which aging and environmental exposures lead to attrition^7,56–58^. Given the abundant evidence linking rare variants or short telomere lengths to reduced survival in IPF^16,59^, these findings suggest that rare and common telomere-associated variants not only underlie genetic heterogeneity, but also drive heterogeneity of disease outcomes.

We find that the overall genetic architecture of IPF is significantly correlated with that of telomere length in contrast to other epidemiologically associated traits. This parallel analysis utilized existing GWAS summary statistics to examine the genetic overlap between these traits and complements our prior Mendelian randomization study implicating telomere shortening as causal for IPF^4^. While genome-wide significant loci from GWAS studies of IPF and telomere length overlap to a minimal degree, the extent of genetic correlation between these two traits suggest that many other variants below significance threshold are also overlapping.

Our findings suggest that the extent of this overlap may be related to genetic heterogeneity of the IPF GWAS cohort. Rare variant carriers, which are enriched in familial pulmonary fibrosis, have attenuated effects from some common variants on disease risk. As such, a sporadic IPF cohort without rare variant carriers could be enriched for polygenic discovery. Similarly, a hypothetical GWAS analysis of an IPF cohort without short telomeres would likely yield different results from an IPF cohort in which all patients have very short telomere lengths. Notably, a previous IPF GWAS^60^ did not identify the genetic locus in *TERT* whereas a similarly powered contemporaneous GWAS did^61^. Imbalance of genetic endotypes may have led to discrepant results in these studies. Our findings highlight the importance of understanding genetic heterogeneity when analyzing genetic association in all-comer IPF cohorts.

Our study has several limitations. Due to the variability in clinical phenotyping of cohorts, some relevant risk factors were not uniformly available including smoking, environmental exposures, and other comorbid conditions. Telomere length genetically correlates with some of these risk factors. Confounding or effect modification from non-genetic factors is not definitively excluded and the impact of gene by environment will require further investigation. Since all major GWAS studies on IPF focused on European ancestry individuals, polygenic scores and associations may not be portable across non-European ancestries. Our sensitivity analyses using non-European cases and controls were limited by sample size and suggested a consistent but non-significant trend for some associations. Future studies with diverse IPF cohorts will be needed to identify pan-ancestry genetic effects. The UKBB IPF cases were relatively depleted of *TERT* and other rare variants compared to clinical cohorts. This may be related to enrollment bias in the UKBB which preferentially recruited healthy older individuals, potentially excluding some individuals with pathogenic rare variants who tend to have younger onset of severe disease. Clinical IPF cohorts may also be biased towards enrolling those with a genetic etiology; a recent analysis of the US-wide Pulmonary Fibrosis Foundation Registry revealed a comparable 7% prevalence of rare variants in IPF cases using the same definition^23^. We grouped all rare variant carriers together due to limited sample size. As such, we were unable to draw definitive conclusions for specific genes, although we observed consistent associations in the most well-represented genes.

In conclusion, polygenic background and rare variation contribute to genetic risk of IPF both independently and in tandem. While attenuated, polygenic modifiers of disease risk exist even amongst IPF carriers of rare damaging variants. We find that genetically predicted short telomeres intersects with IPF risk not only in the purview of damaging rare variants, but also in the context of polygenic risk in certain endotypes. Accounting for these novel genetic risk factors both improves disease prediction and explains additional genetic liability. Taken together, our findings underscore the importance of understanding IPF heterogeneity from rare and common genetic influences that may be relevant for future studies.

## Contributors

DZ conceived the project and designed the study. DZ, AD, LJ, HG, AC, and GH performed the analysis. AK, CW, CKG and KK generated genetic data for the discovery cohort. CKG, IN, FJM, GR, CAN, and the Columbia Genomics Consortium enrolled and contributed samples for the discovery cohort. MM and MC provided analytic support for alternative polygenic scores. MG and CJS provided supervisory support to AD and facilitated access to UK Biobank. DZ, AD, and CKG interpreted the results and drafted the manuscript.

## Data sharing

Genotype data for discovery cohort allowable under consent is available in the Database of Genotypes and Phenotypes (dbGAP) under the following projects: Pulmonary Fibrosis and Telomerase Dysfunction, phs002692; Genomics of Glomerular Disorders, phs002480; Genomic Translation for ALS Care (GTAC), phs02973. The TOPMed cohort genetic and phenotypic data are available under dbGAP (Multi-ethnic Study of Atherosclerosis, phs0001416; Framingham Heart Study, phs000974; Idiopathic Pulmonary Fibrosis, phs001607). Genetic and phenotypic data from the UK Biobank is available through applications to the UK Biobank.

## Acknowledgements

The authors thank the research participants for their participation and enrollment in study cohorts. The authors thank the investigators and institutions who supported the NHLBI cohorts (IPF, MESA, and the Framingham Heart Study). The authors would like to acknowledge the Collaborative Group of Genetic Studies of IPF for access to summary statistics of prior IPF GWAS. The research utilized data from the UK Biobank resource carried out under UK Biobank application number 103356. UK Biobank protocols were approved by the National Research Ethics Service Committee. This study was funded by the National Institute for Health and Care Research Exeter Biomedical Research Centre. The views expressed are those of the author(s) and not necessarily those of the NIHR or the Department of Health and Social Care. The <u>Columbia Genomics Consortium</u> includes data and investigators involved with the following projects: Cure Glomerulonephropathy (CureGN, www.cureGN.org; Ali Gharavi, Krzysztof Kiryluk, and Simone Sanna-Cherchi), non-cureGN (Ali Gharavi, Krzysztof Kiryluk, Simone Sanna-Cherchi), Genomic Translation for ALS Care (GTAC; Matthew Harms and David Goldstein), and Columbia University Biobank cohorts for COVID-19 (Krzysztof Kiryluk, Muredach Reilly, Soumitra Sengupta, Eldad Hod, Wendy Chung, Jennifer Williamson), atopic disease (Joshua Milner and Jordan Orange), prenatal disease (Ronald Wapner), and diagnostic sequencing (Muredach Reilly) The “Genomics of Glomerular Disorders” project (dbGAP phs002480) includes genetic data for the participants of the CureGN Study (www.curegn.org) supported by the National Institute of Health (NIH)/National Institute of Diabetes and Digestive and Kidney Diseases (NIDDK). The sequence data was generated in collaboration with AstraZeneca as part of the High Impact, Interdisciplinary Science Grant RC2-DK116690 from the NIH/NIDDK. The “Genomic Translation for ALS Care” project (dbGAP phs020973) includes the following investigators and funding sources: PI Matthew Harms (Columbia University) and site investigators Stanley Appel (Houston Methodist); Robert Baloh (Cedars Sinai); Richard Bedlack (Duke Univ); Siddharthan Chandran (Univ. of Edinburgh); Laura Foster (Univ of Colorado); Summer Gibson, (Univ Utah); David Goldstein (Columbia University); Stephen A. Goutman (Univ of Michigan); Chafic Karam (Oregon Health Sciences); David Lacomis (Univ. of Pittsburgh); George Manousakis (Univ. of Minnesota); Timothy Miller (Washington Univ St. Louis); Cristiane Moreno (Columbia University); Suvankar Pal (Univ. of Edinburgh); Dhruv Sareen (Cedars Sinai); Zachary Simmons (Penn State Hershey); Leo Wang (Univ of Washington). Funding was provided by the ALS Association (National); ALS Association (Greater New York Chapter), and Biogen. The “Pulmonary Fibrosis and Telomerase Dysfunction” project (dbGAP phs002692) was supported by grant HL093096 from NIH/NHLBI to Christine Kim Garcia (Columbia University) with assistance from David Goldstein (Columbia University) and Chad A. Newton (UT Southwestern).

