## Supplementary material for "Polygenic Risk and Rare Variants in Endotypes of Idiopathic Pulmonary Fibrosis": online supplement

### Methods


**Figure S1.** Hudson plot of genetic association studies of IPF and telomere length.

**Figure S2**. Principal components and genetic ancestry.

**Figure S3.** Distributions of polygenic scores in IPF and controls.

**Figure S4**. Distributions of polygenic scores in IPF rare variant carriers and non-carriers.

**Figure S5.** Distributions of polygenic scores in IPF and controls in non-Europeans.

**Figure S6.** Distributions of polygenic scores in endotypes of IPF in the Columbia cohort.

**Figure S7**. Significance of pairwise interaction terms of genetic variables

**Figure S8.** Sensitivity analyses for associations with IPF risk using alternative polygenic scores.

**Figure S9.** Association of polygenic scores with IPF risk by rare variant carrier status in the TOPMed cohort.

**Figure S10.** Association of polygenic scores with IPF risk in European-only ancestry individuals.

**Figure S11.** Association of polygenic scores with IPF risk in Non-European ancestry individuals.

**Figure S12.** Association of polygenic scores with IPF risk in rare variant carriers by specific gene.

**Figure S13**. Proportion of liability of IPF explained by genetic variables in general population for non-European ancestry individuals.

**Figure S14**. Proportion of liability of IPF explained by polygenic scores using alternative 20-SNP telomere length polygenic score in the general population.

**Figure S15**. Proportion of liability of IPF explained by polygenic scores using alternative lasso regression IPF polygenic score in the general population.

**Figure S16.** Pair-wise genetic correlation amongst IPF and epidemiologically associated traits.

**Figure S17**. Genetic correlation between IPF and epidemiologically associated traits.

**Table S1.** Genome-wide significant SNPs associated with IPF

**Table S2.** Genome-wide significant autosomal SNPs associated with telomere length

**Table S3.** Overlapping genetic loci in GWAS studies of IPF and telomere length

**Table S4.** Descriptions of polygenic scores

**Table S5.** Phenotypes of controls in Columbia discovery cohort.

**Table S6.** Age, sex, and ancestry of cohort participants.

**Table S7.** Genes represented by rare variant carriers in IPF cohorts.

**Table S8.** Characteristics of IPF endotypes in all cohorts.

### Methods

*Patient cohorts*

The Columbia IPF cohort with whole genome sequencing (WGS) data has been previously described[1,2]. Briefly, the institutional review board at Columbia University Medical Center (CUMC; #AAAS0753 and #AAAS7495) approved this study. Patients with pulmonary fibrosis were collected by CUMC, the University of Texas Southwestern, and the IPF Clinical Research Network PANTHER-IPF[3] (Evaluating the Effectiveness of Prednisone, Azathioprine, and N-acetylcysteine in Patients With IPF; NCT00650091) and ACE-IPF[4] (Anticoagulant Effectiveness in Idiopathic Pulmonary Fibrosis; NCT 00957242) clinical trials including only those individuals who consented to participate in both the parent study and the optional genetic substudy. Patients with IPF or familial pulmonary fibrosis at recruitment at CUMC or UT Southwestern were enrolled in the original cohort; over time longitudinal assessment resulted in diagnostic change due to clinical manifestations or guideline revisions. Only patients meeting IPF diagnosis by current guidelines were included for this study. Non-IPF disease controls with whole genome sequencing data sequenced and processed under the same pipeline at CUMC were available for comparison as previously described **(Table S5)**[2].

The TOPMed cohort included WGS data made available with permission through the database of Genotypes and Phenotypes (dbGaP) as previously described[2]. Briefly, a cohort of IPF subjects (phs001607) and control subjects from the Multi-Ethnic Study of Atherosclerosis (MESA; phs001416) and the Framingham Heart Study (FHS, phs000974) were used. Available phenotypes including age and sex were accessed through dbGaP. IPF cases and controls were pruned for relatedness or duplicate samples both within the TOPMed cohort and between the TOPMed and the Columbia cohorts.

The UK Biobank cohort with WGS data and available clinical and molecular phenotypes has been previously described[5]. Briefly, cases were defined using coded electronic diagnoses including hospital episode statistics, primary care records and death records (ICD10 code J84.1 for cases and exclusion of all J84 for controls). In some UKBB cases, diagnosis of IPF was made after study registration and blood draw for telomere length measurement. We used age of registration for adjustment but also reported age of diagnosis.

All cohorts were restricted to unrelated subjects with available genotypes, age, and sex **(Figure 1A, Table S6)**.

*Genome Sequencing and Variant Calling*

Whole genome sequencing was performed at Columbia University using standard protocols on Illumina's NovaSeq 6000 platform. Illumina lane-level FASTQ files were aligned to Human Reference Genome GrCh37 using DRAGEN. Sample-level BAM files were jointly genotyped following Genome Analysis Toolkit (GATK)[6] best practices and using GATK tools to obtain rare and common variant genotypes. Site-level QC was performed to exclude variants failing GATK's variant quality score recalibration (VQSR) filters. Low-quality individual genotypes were set as missing if read depth (DP) < 10, genotype quality (GQ) < 20, or allelic balance fell outside of 0.2-0.8 for heterozygous calls. Variant Effect Predictor (VEP)[7] and dbNSFP[8] were used to annotate variants with predicted function, gnomAD[9] allele frequencies, and in silico tools including REVEL[10] and PrimateAI[11]. Rare qualifying variants in disease-associated genes predicted to be deleterious to resultant gene product were identified using above *in silico* tools as previously described[1,2]. *KING*[12] was used to estimated kinship and those with more than third-degree relatedness were excluded. Principal components used for adjustment of population substructure was obtained using *plink*[13] after pruning variants for linkage disequilibrium. Details and use of the TOPMed WGS cohort[14] and the UKBB[5] WGS 450k cohort has been described previously.

*Ancestry Prediction*

For the Columbia and TOPMed cohorts, we used *peddy*[15] to infer genetic ancestry of subjects using a machine learning model to sample 25,000 variants and perform principal component analysis to calibrate ancestry against 1000 Genomes reference data[16]. We used default prediction cutoffs of 95% to assign ancestry.

For the UKBB cohort, genetic ancestry was computed using principal component analysis compared against self-reported ethnic background as previously described[5].

*Rare Qualifying Variant Definitions*

Detailed definitions of bioinformatic filters used for inferring rare deleterious qualifying variants in IPF-associated genes (*TERT, TERC, RTEL1, PARN, DKC1, TINF2, NAF1, ZCCHC8, SFTPC, SFTPA1, SFPTA2, KIF15*)[1] have been previously described[2]. We updated definitions using gnomAD[9] v4 for population allele frequencies. Rare qualifying variants were included if they were below population allele frequencies of 0.0005 and predicted to be loss of function mutations (eg. frameshift, stop gained, start lost), in-frame insertion-deletions, or missense mutations passing majority *in silico* filters (Polyphen2[17] probably damaging, REVEL[10] score ≥ 0.5, and PrimateAI[11] score ≥ 0.8). As before, we included rare *TERC* variants if they had a population maximum allele frequency below 0.0005 and disrupted intramolecular base-paring.

*Polygenic scores*

We extracted genome-wide significant SNPs from published GWAS studies of IPF and telomere length to generate polygenic scores. The Columbia discovery cohort samples were not included in GWAS cohorts[18,19] used for generating the IPF-PRS-no*MUC5B*. The TL-PRS was generated from UKBB samples by their association with telomere length, not IPF. Due to the outsized effect and prevalence of the *MUC5B* rs35705950 promoter polymorphism, we analyzed this variant separately from the IPF polygenic score as previously described. Three genome-wide significant loci were in linkage disequilibrium in the GWAS studies for IPF and telomere length used for polygenic scores **(Figure S1)**; two of the three loci are the top significant SNPs associated with telomere length **(Table S3)**. We compared raw distributions of the telomere length polygenic score (TL-PRS) and different versions of the IPF polygenic score including *MUC5B* (IPF-PRS-*MUC5B*), excluding *MUC5B* but retaining the overlapping telomere-associated loci (IPF-PRS-no*MUC5B*-withTelo) and excluding both *MUC5B* SNP and the overlapping telomere-associated loci (IPF-PRS-no*MUC5B*-noTelo). We also explored alternative polygenic scores for IPF using a previously described LASSO regression approach[20] that excludes 500 kbp flanking the *MUC5B* SNP (IPF-PRS-lasso), a telomere polygenic score that excludes the shared telomere-associated loci (TL-PRS-noTelo), and a telomere length polygenic score using a different GWAS study[21] that yielded 20 genome-wide significant SNPs (TL-PRS-20snp) **(Table S4)**. For primary analysis, we used non-overlapping polygenic scores and focused on the TL-PRS (referred to as ***"TL-PRS"*** in the main manuscript) and the IPF-PRS-no*MUC5B*-noTelo (referred to as ***"IPF-PRS-noMUC5B"*** in the main manuscript). We explored alternative polygenic scores including the IPF-PRS-no*MUC5B*, IPF-PRS-lasso, TL-PRS-noTelo, and TL-PRS-20snp for sensitivity analyses **(Figure S6).**

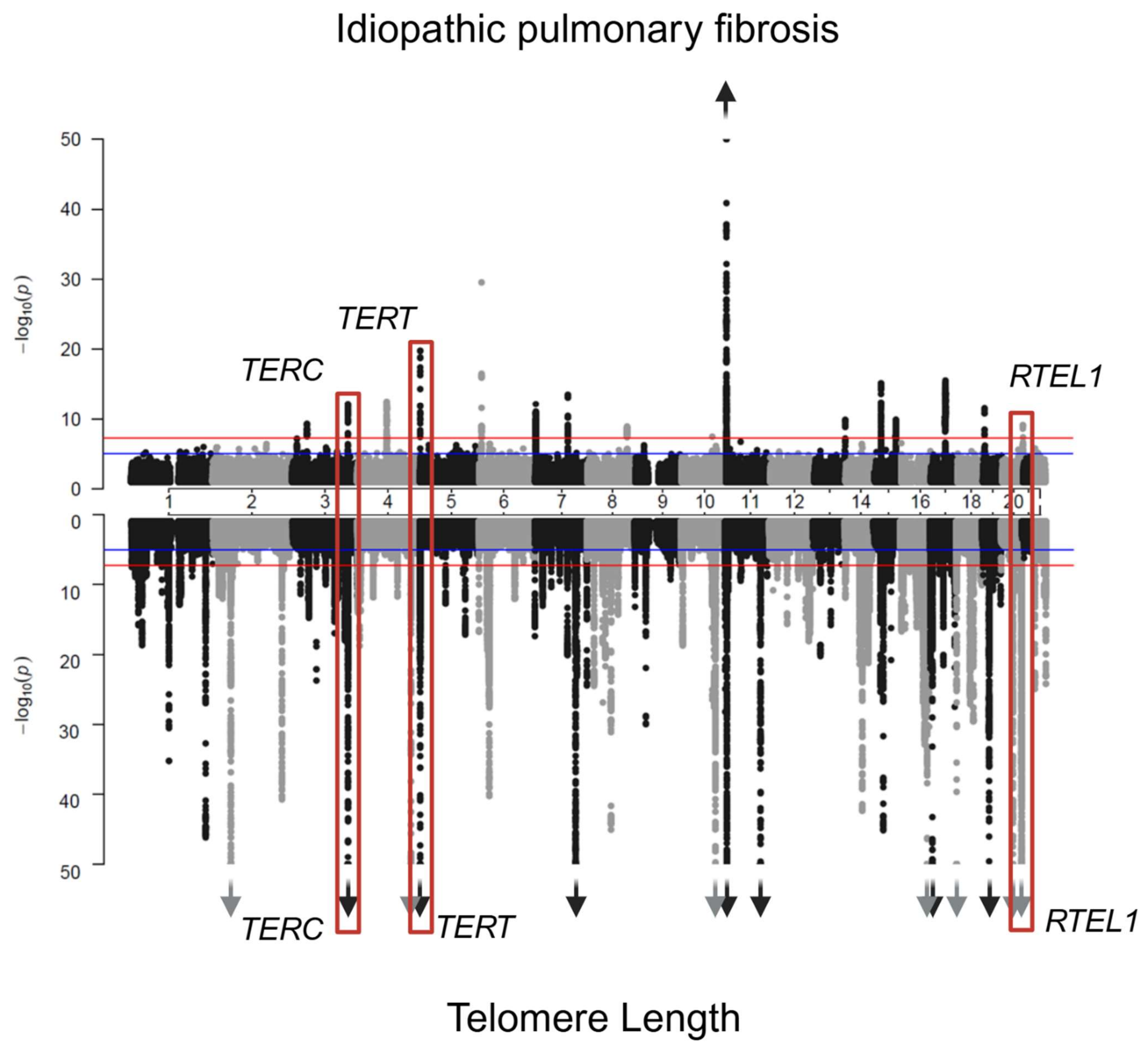


**Supplemental Figure 1**. **Hudson plot of genetic association studies of IPF and telomere length**. Summary statistics of published GWAS studies of IPF[18] and telomere length[19] shown. For ease of visualization, the y-axis is truncated at negative log p-value of 50 for both studies with arrows indicating loci that exceed this threshold. Three loci shown in red are in linkage disequilibrium between both studies highlighting overlap of genome-wide significant signals related to telomere length and IPF risk.

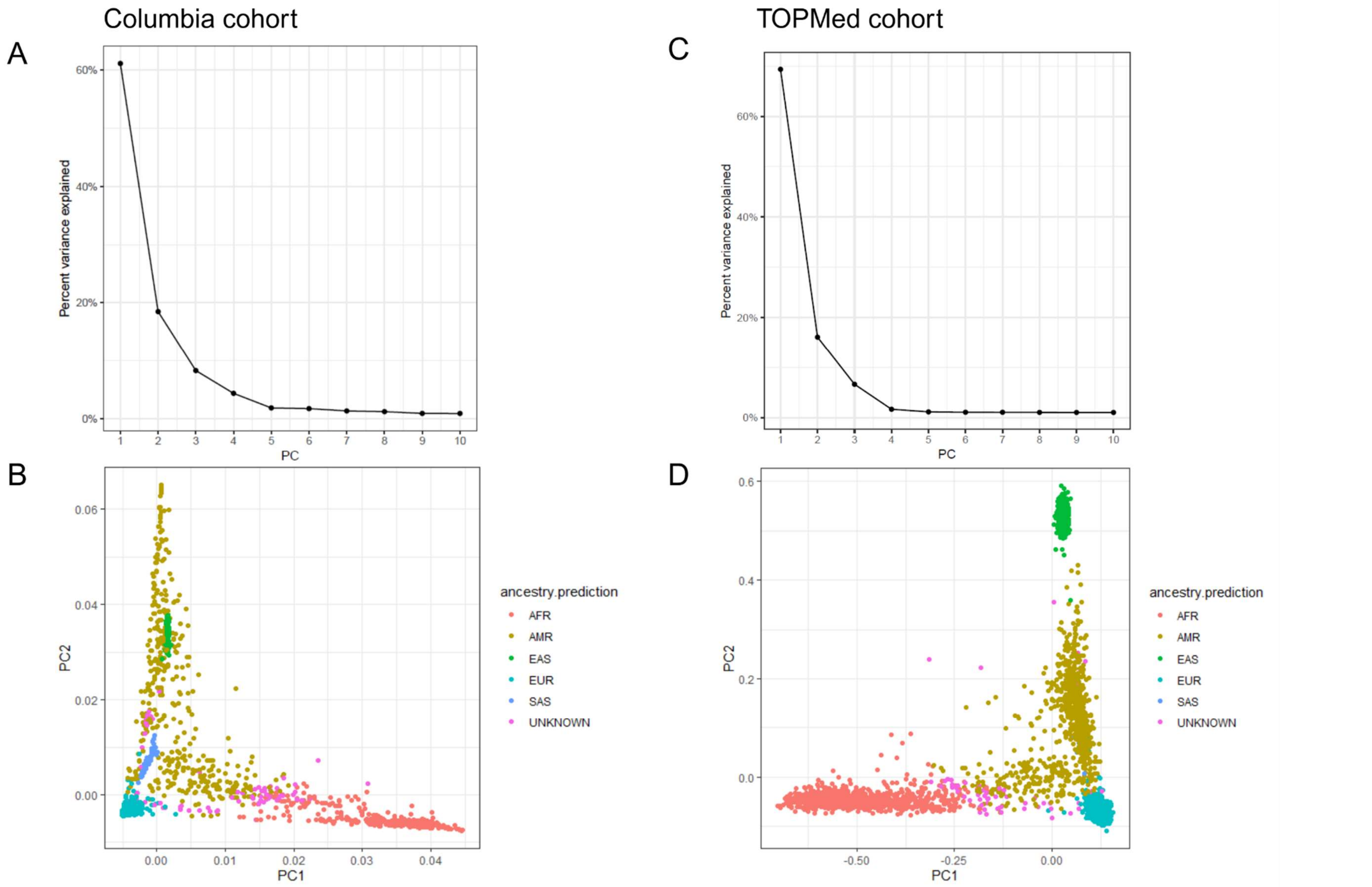


**Supplemental Figure 2**. **Principal components and genetic ancestry**. Percent of variance explained by first ten principal components visualized for the Columbia cohort **(A)** and TOPMed cohort **(B)**. Overlay of genetic ancestry predicted by peddy[15] and first two genetic principal components shown for Columbia cohort **(C)** and TOPMed cohort **(D)**.

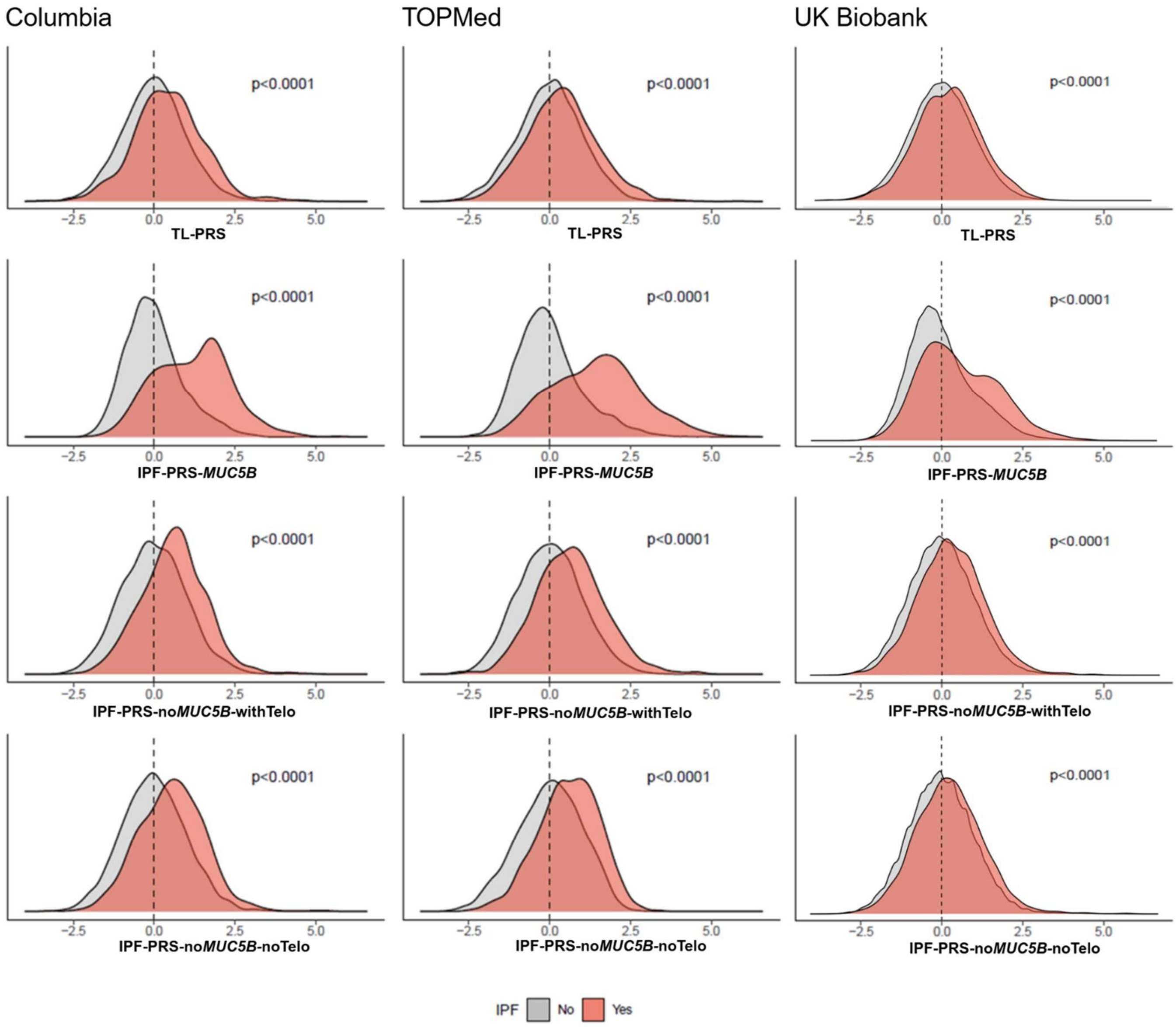


**Supplemental Figure 3**. **Distributions of polygenic scores in IPF and controls**. P-values indicate results of Wilcoxon rank sum test comparing polygenic scores in cases versus controls. IPF polygenic scores are displayed including (**IPF-PRS-*MUC5B***), excluding the MUC5B *rs35705950* promoter polymorphism (**IPF-PRS-no*MUC5B*-withTelo**), and excluding overlapping telomere-associated genetic loci (**IPF-PRS-no*MUC5B*-noTelo**). All polygenic scores are displayed as control-normalized Z-transformed scores.

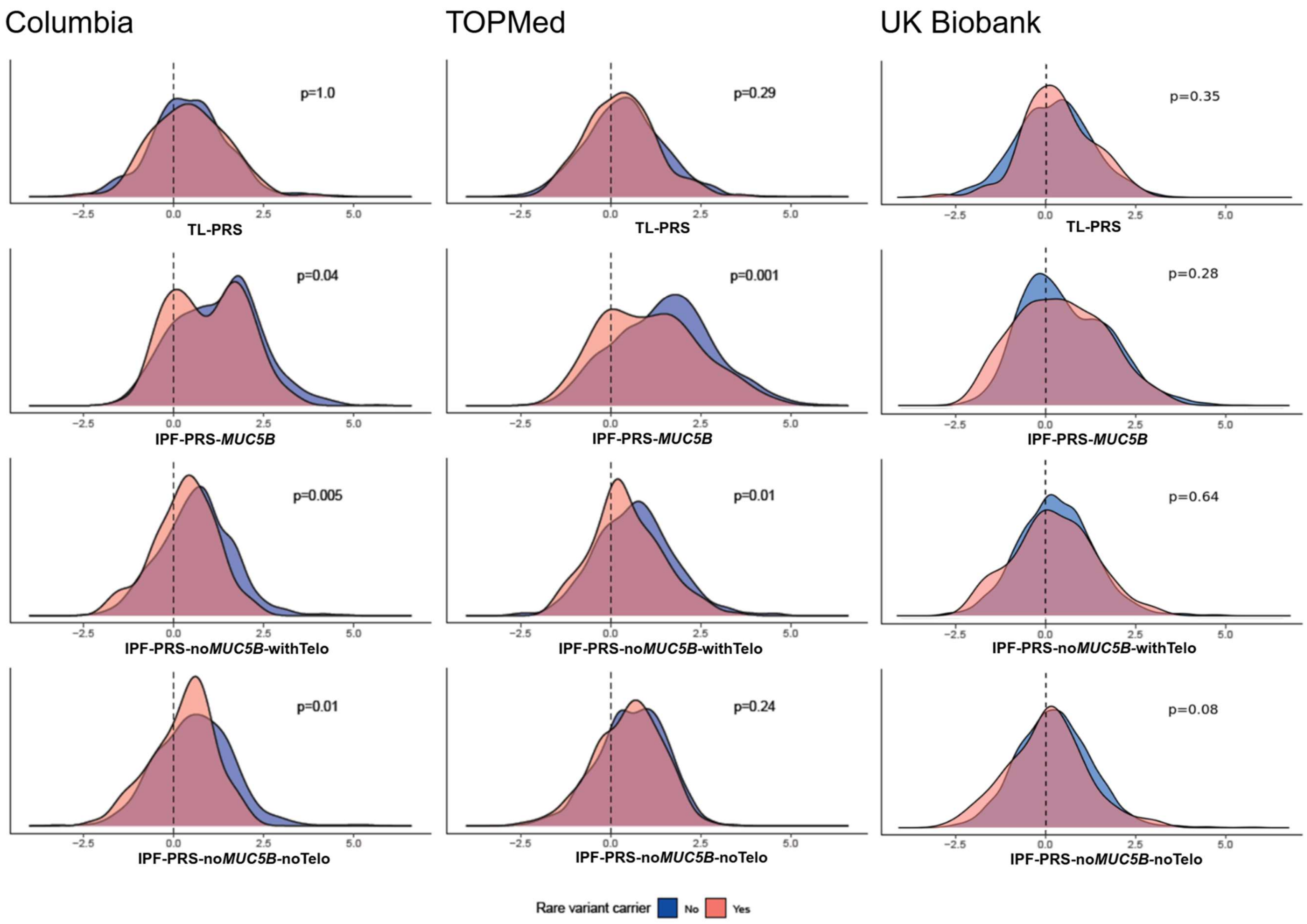


**Supplemental Figure 4**. **Polygenic scores in IPF rare variant carriers and non-carriers**. P-values indicate results of Wilcoxon rank sum comparing IPF rare variant carriers versus IPF non-carriers. IPF polygenic scores are displayed including (**IPF-PRS-*MUC5B***), excluding the MUC5B *rs35705950* promoter polymorphism (**IPF-PRS-no*MUC5B*-withTelo**), and excluding overlapping telomere-associated genetic loci (**IPF-PRS-no*MUC5B*-noTelo**). All polygenic scores are displayed as control-normalized Z-transformed scores (controls not displayed).

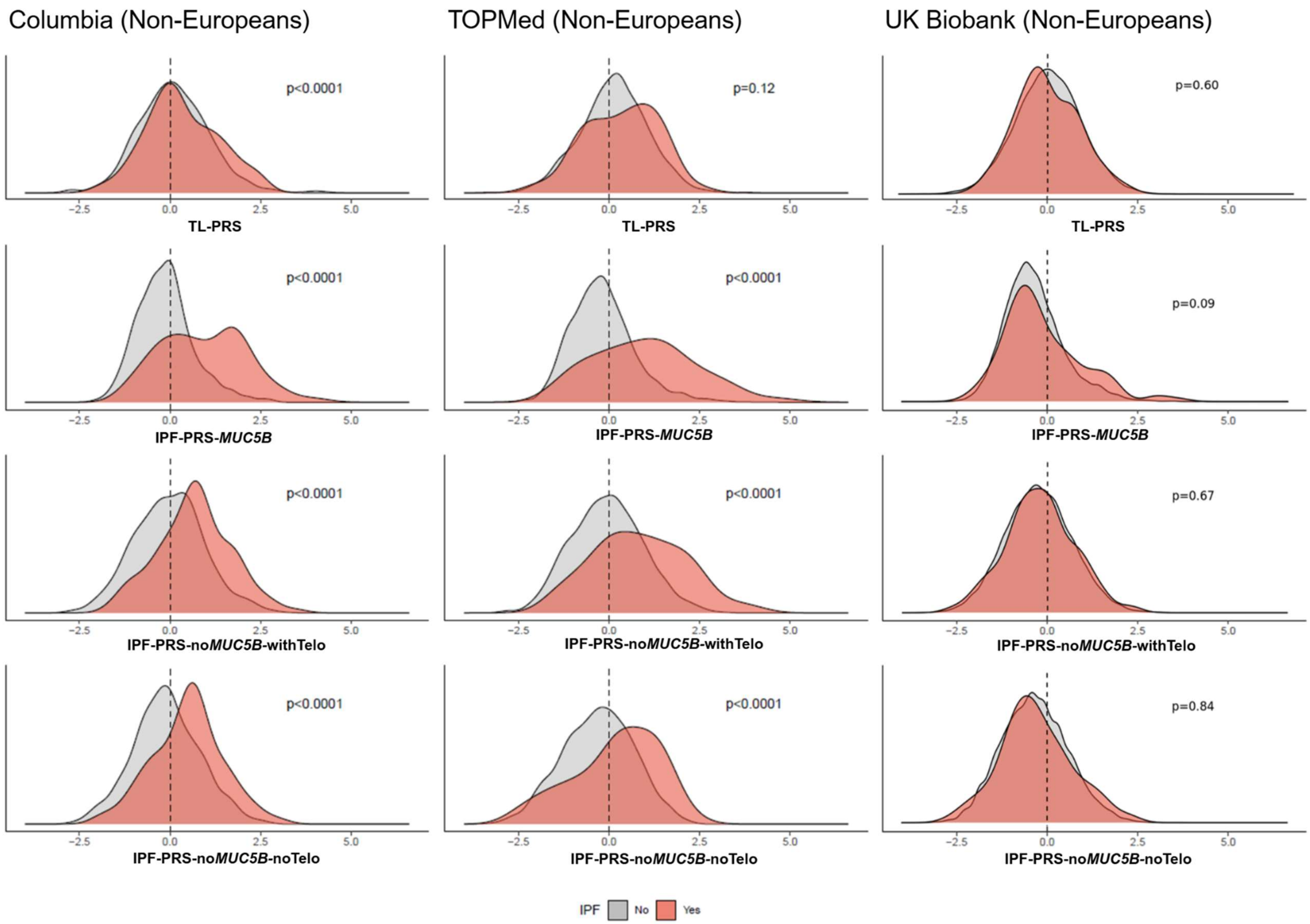


**Supplemental Figure 5**. **Distributions of polygenic scores in IPF and controls in non-Europeans**. P-values indicate results of Wilcoxon rank sum test comparing polygenic scores in cases versus controls of non- European genetic ancestry. IPF polygenic scores are displayed including (**IPF-PRS-*MUC5B***), excluding the MUC5B *rs35705950* promoter polymorphism (**IPF-PRS-no*MUC5B*-withTelo**), and excluding overlapping telomere-associated genetic loci (**IPF-PRS-no*MUC5B*-noTelo**).All polygenic scores are displayed as control-normalized Z-transformed scores.

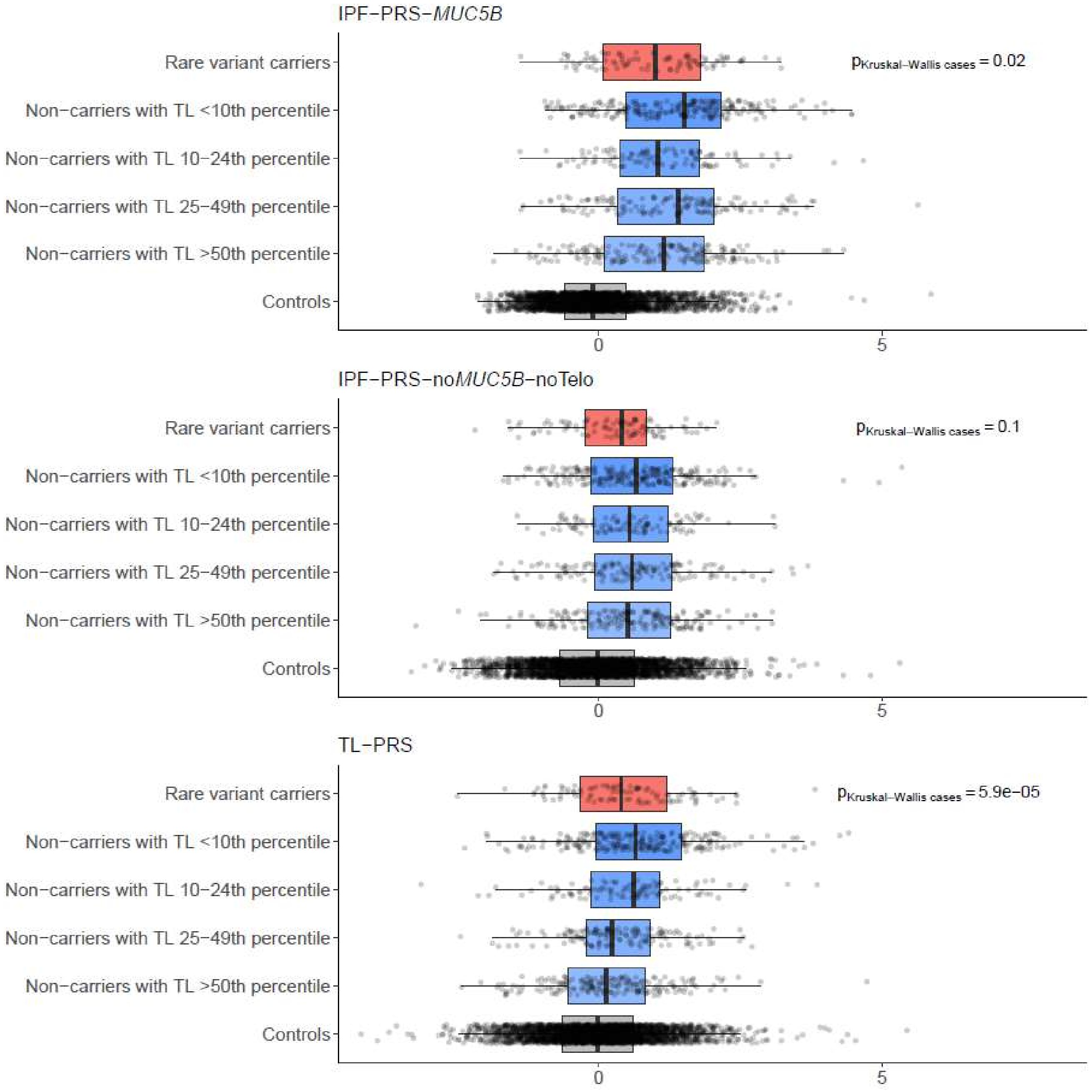


**Supplemental Figure 6**. **Distributions of polygenic scores in endotypes of IPF in the Columbia cohort**. Boxplot shows control-normalized Z-transformed polygenic scores across different endotypes of IPF stratified by rare variant carrier status and telomere length strata. P-values indicate results of Kruskal-Wallis test in cases to assess heterogeneity of polygenic scores amongst IPF endotypes. IPF polygenic scores are displayed including (**IPF-PRS-*MUC5B***) and excluding both the *MUC5B rs35705950* promoter polymorphism and overlapping telomere-associated genetic loci (**IPF-PRS-no*MUC5B*-noTelo**).

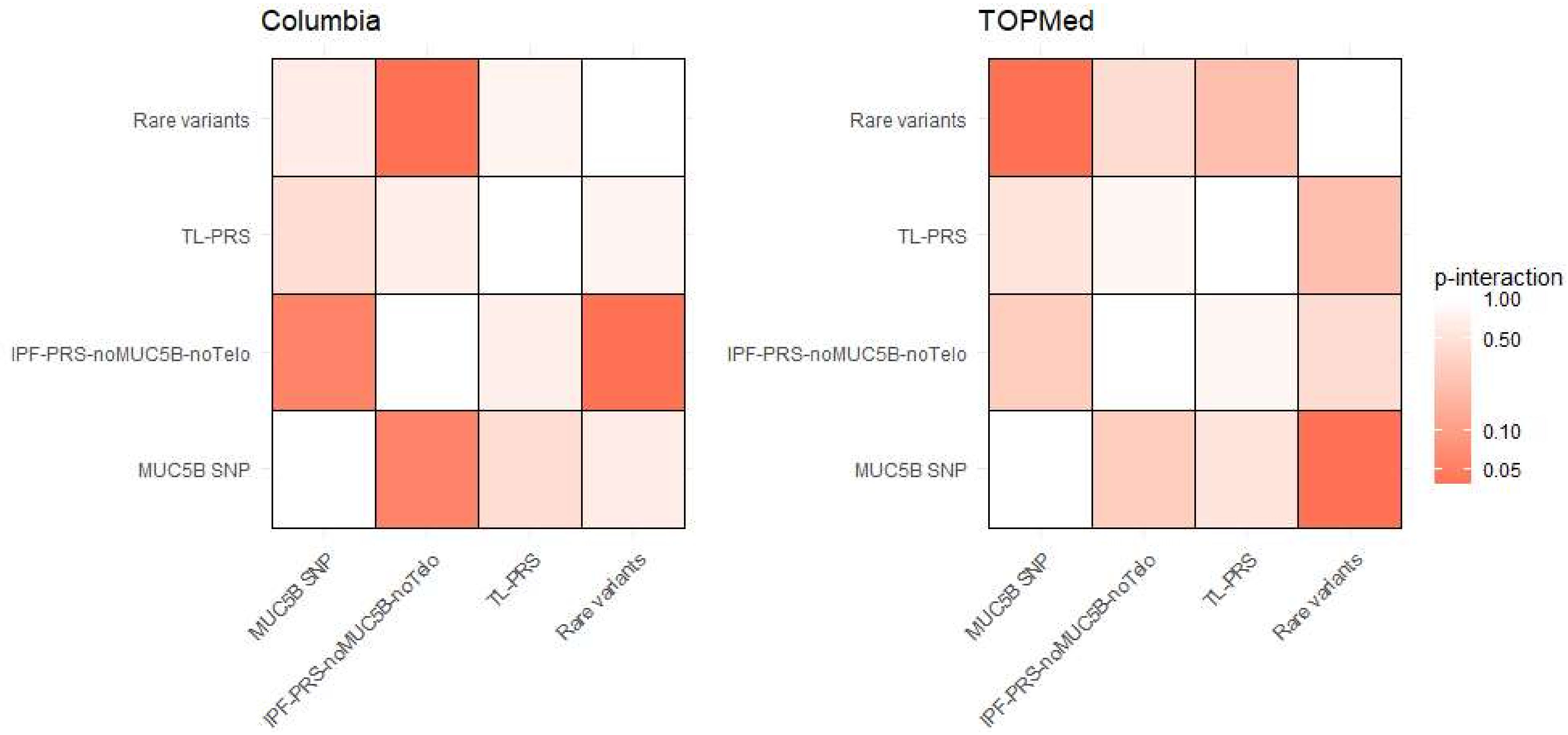


**Supplemental Figure 7**. **Significance of pairwise interaction terms of genetic variables.** For each pair of genetic variables, an interaction term was tested along with all other individual genetic variables, age, sex, and 5 PC of ancestry in a multivariable logistic regression model with IPF status as outcome. No pair-wise interaction was significant after correcting for multiple comparisons. IPF-PRS-no*MUC5B*-noTelo is computed excluding the *MUC5B* rs35705950 polymorphism and the three shared genetic loci associated with telomere length.

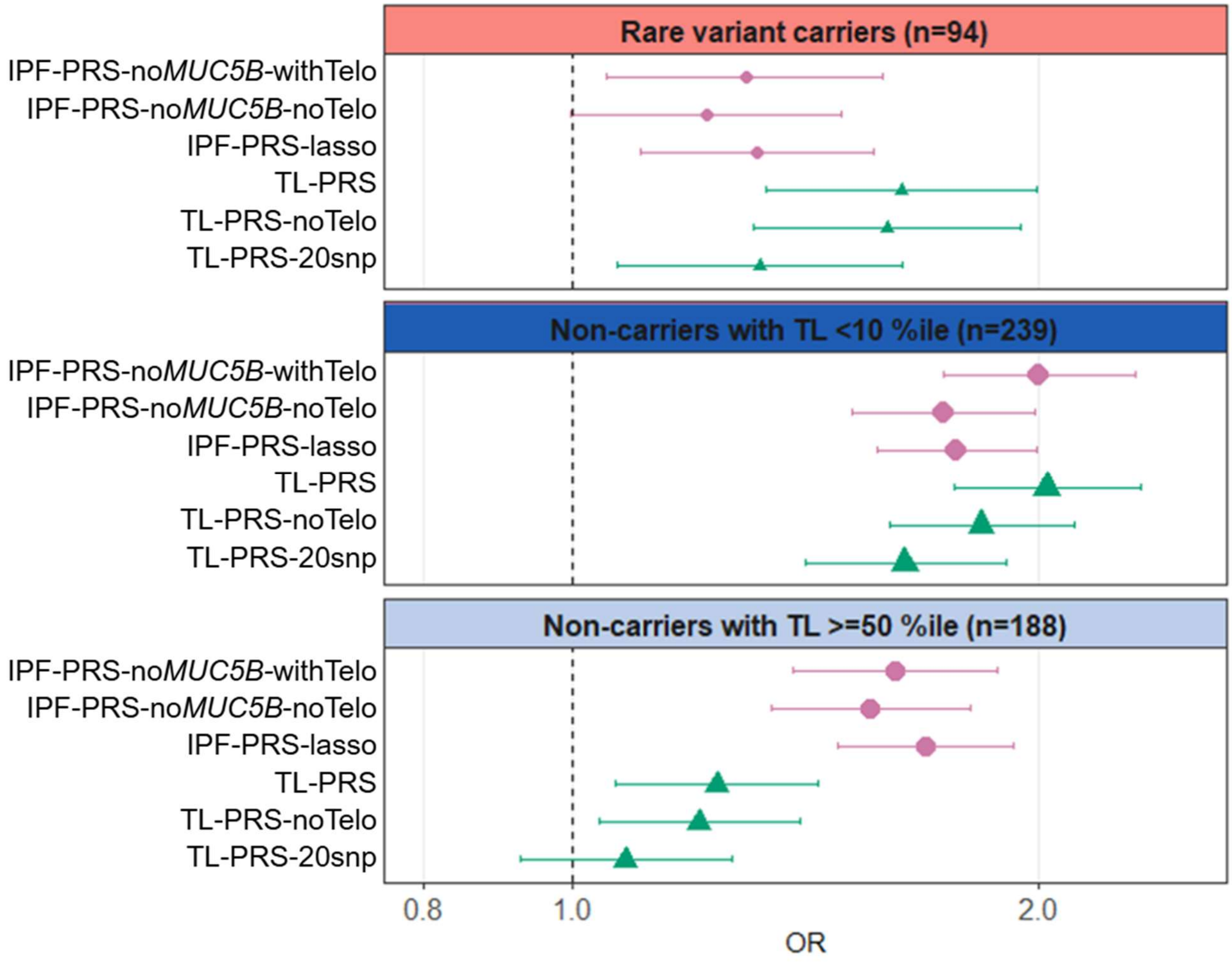


**Supplemental Figure 8**. **Sensitivity analyses for associations with IPF risk using alternative polygenic scores.** Odds ratios and 95% confidence intervals shown using data from Columbia cohort. Associations are adjusted for age, sex, and 5 PC of ancestry. Selected IPF endotypes highlighted to represent diversity in contributions of each polygenic score. IPF polygenic score computed by excluding the *MUC5B* SNP (**IPF-PRS-no*MUC5B*-withTelo**), excluding both the *MUC5B* SNP and the three shared genetic loci associated with telomere length (**IPF-PRS-no*MUC5B*-noTelo**), or using an alternative LASSO method which excludes a 500kb region flanking the *MUC5B* SNP and includes the three shared genetic loci (**IPF-PRS-lasso**).Telomere length polygenic score computed using the full score (**TL-PRS**), excluding the three shared genetic loci (**TL-PRS-noTelo**), or using an alternative telomere length GWAS with 20 genome-wide significant SNPs (**TL-PRS-20snp**).

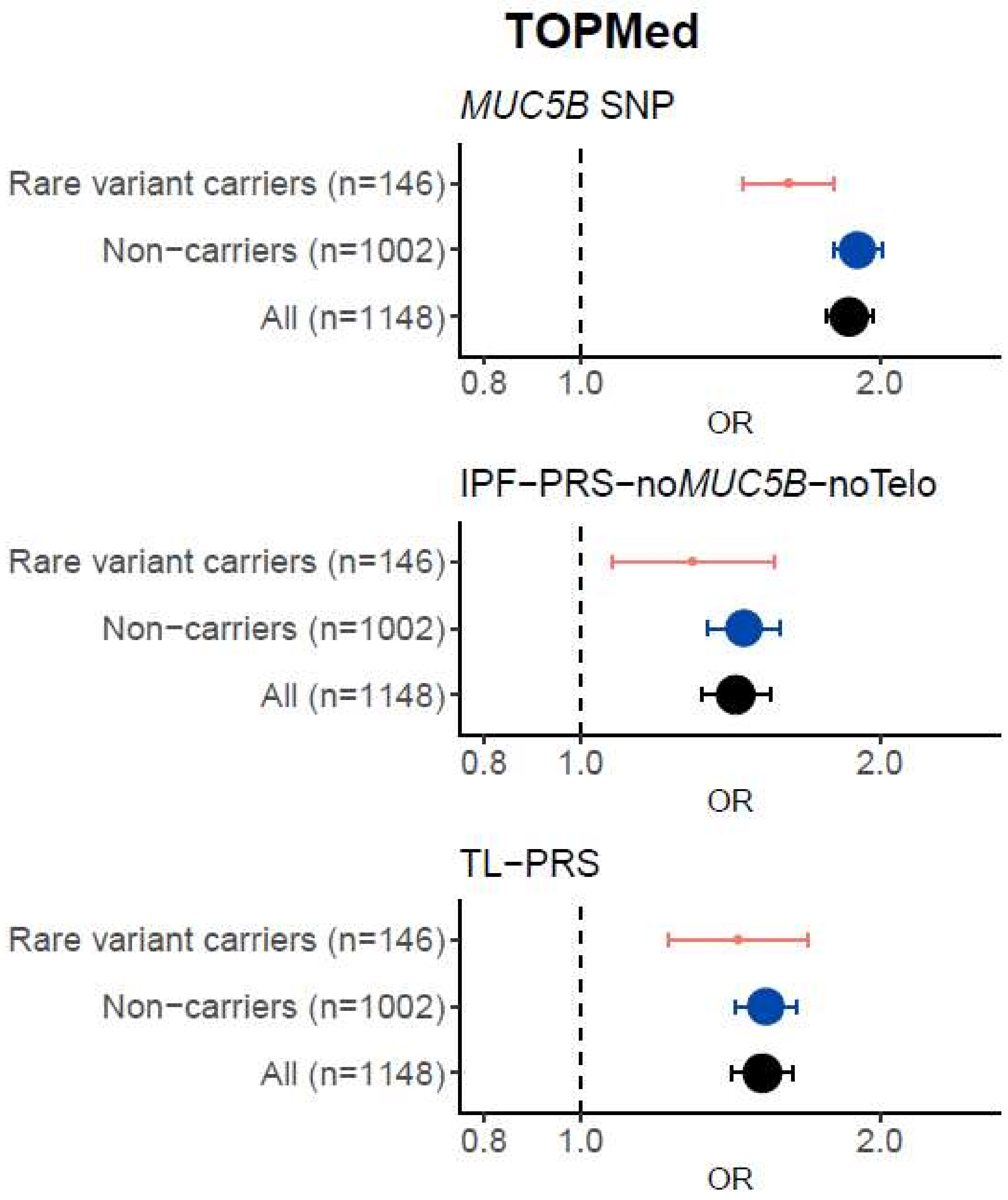


**Supplemental Figure 9**. **Association of polygenic scores with IPF risk by rare variant carrier status in the TOPMed cohort**. Odds ratios and 95% confidence intervals shown for IPF rare variant carriers and non-carriers. All associations with polygenic scores are adjusted for age, sex, and 5 PC of ancestry. IPF-PRS-no*MUC5B*-noTelo is computed excluding the *MUC5B* rs35705950 polymorphism and the three shared genetic loci associated with telomere length.

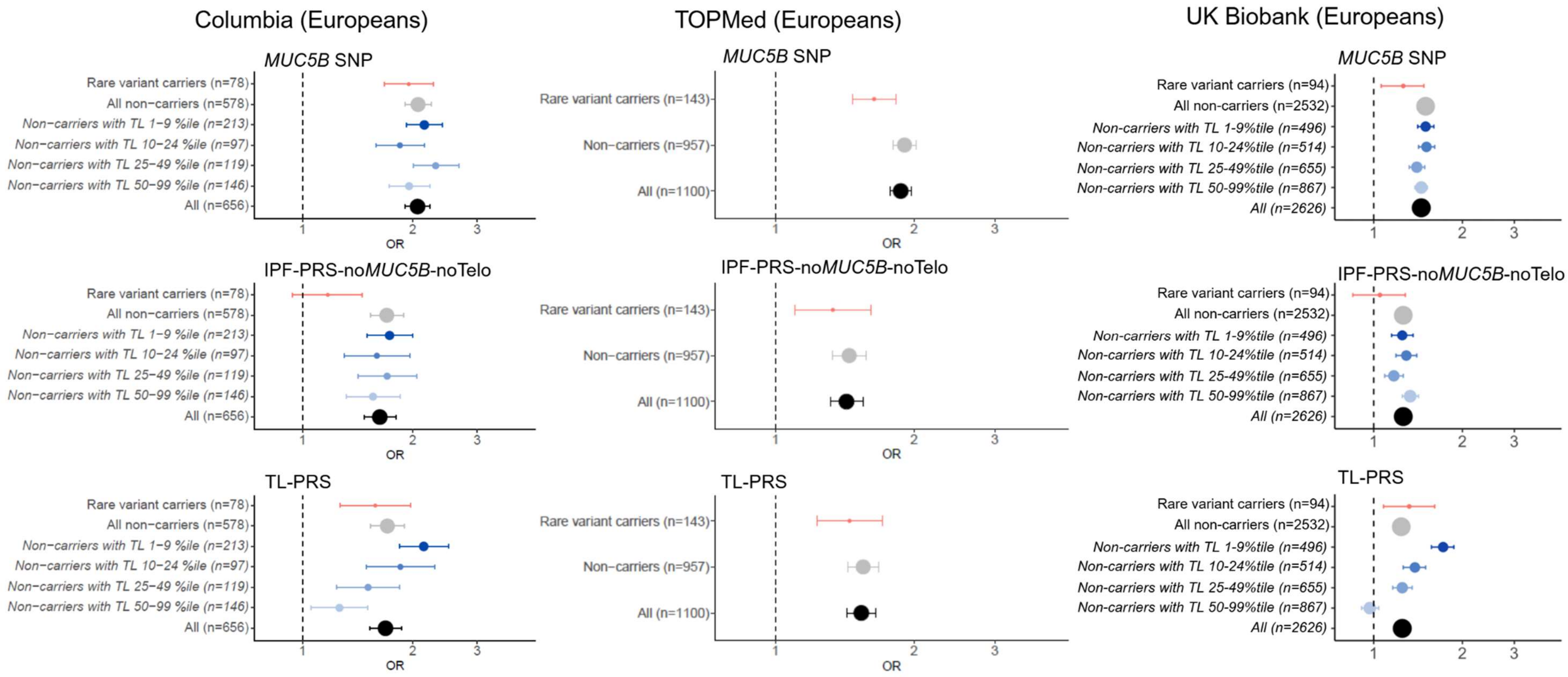


**Supplemental Figure 10**. **Association of polygenic scores with IPF risk in European-only ancestry individuals**. Odds ratios and 95% confidence intervals shown using European-only cases and controls from Columbia discovery cohort, and the TOPMed and UK Biobank replication cohorts. Associations are adjusted for age, sex, and 5 PC of ancestry. IPF-PRS-no*MUC5B*-noTelo is computed excluding the *MUC5B* rs35705950 polymorphism and the three overlapping telomere-associated loci.

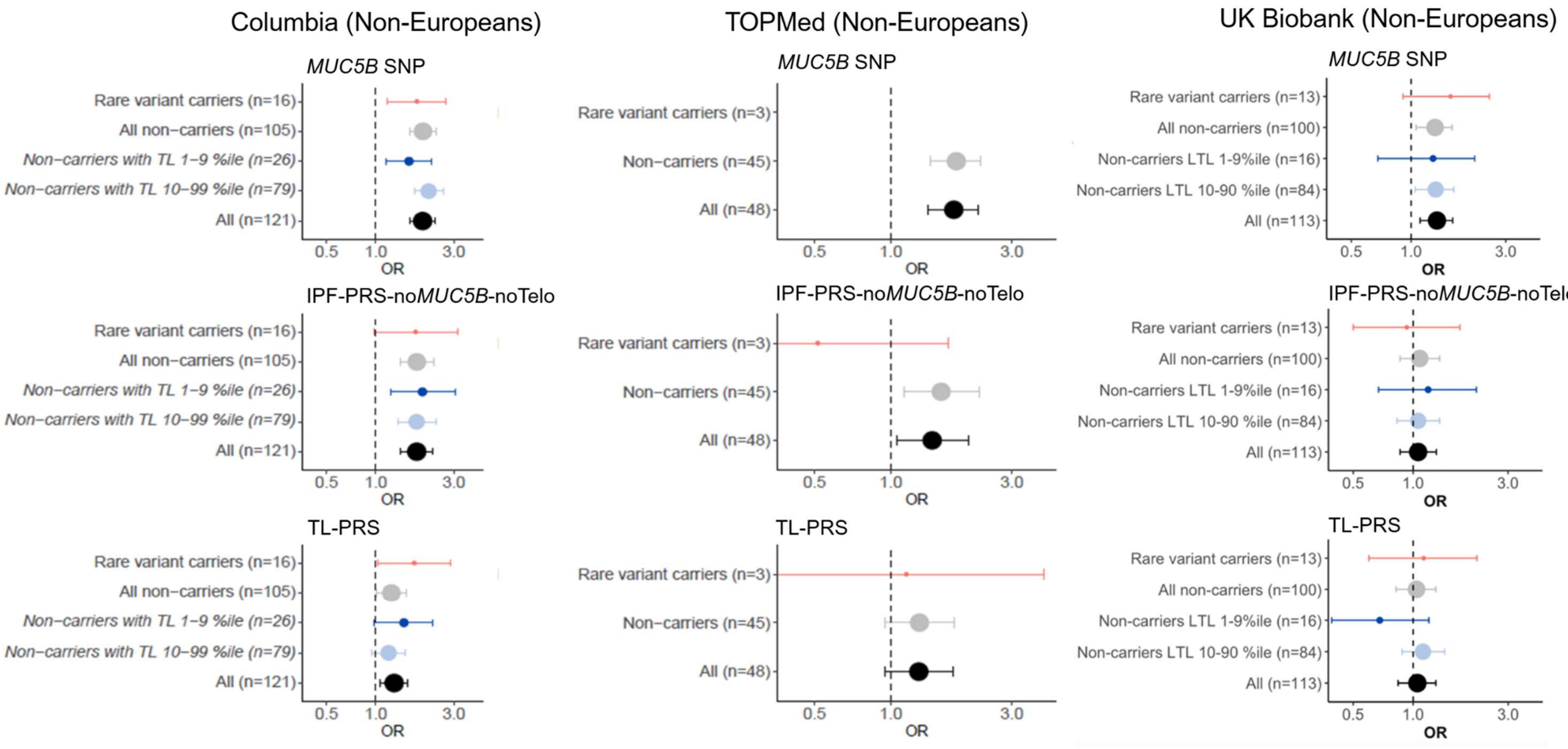


**Supplemental Figure 11**. **Association of polygenic scores with IPF risk in Non-European ancestry individuals**. Odds ratios and 95% confidence intervals shown using non-European cases and controls from Columbia discovery cohort, and the TOPMed and UK Biobank replication cohorts. Associations are adjusted for age, sex, and 5 PC of ancestry. IPF-PRS-no*MUC5B*-noTelo is computed excluding the *MUC5B* rs35705950 polymorphism and the three overlapping telomere-associated loci.

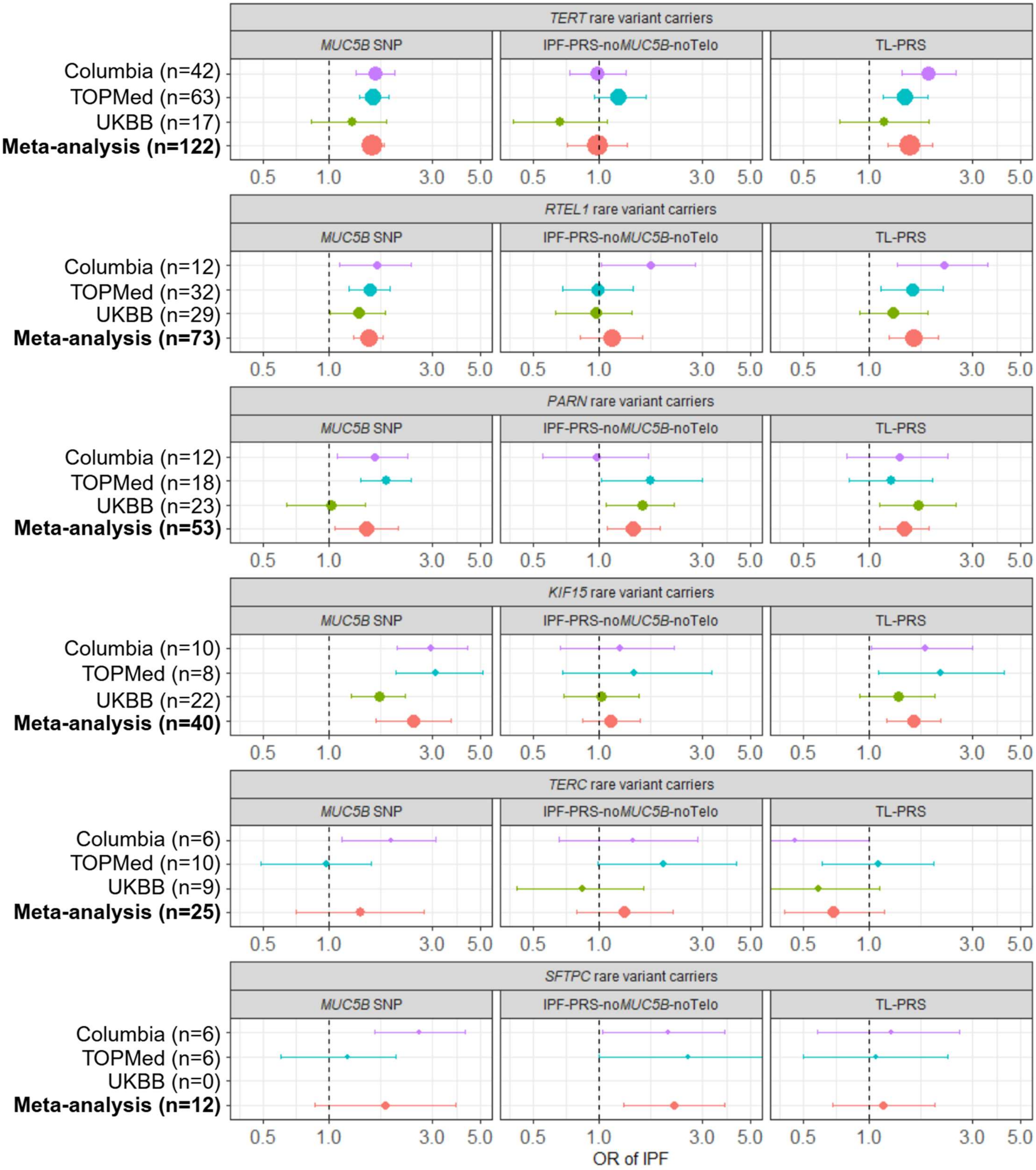


**Supplemental Figure 12**. **Association of polygenic scores with IPF risk in rare variant carriers by specific gene**. Odds ratios and 95% confidence intervals shown using data from Columbia cohort, TOPMed cohort, and UK Biobank. Meta-analysis performed using random-effects model. Associations are adjusted for 2 PC of ancestry due to limited sample size. No *TERC* carriers with IPF in the UKBB had the *MUC5B* polymorphism.

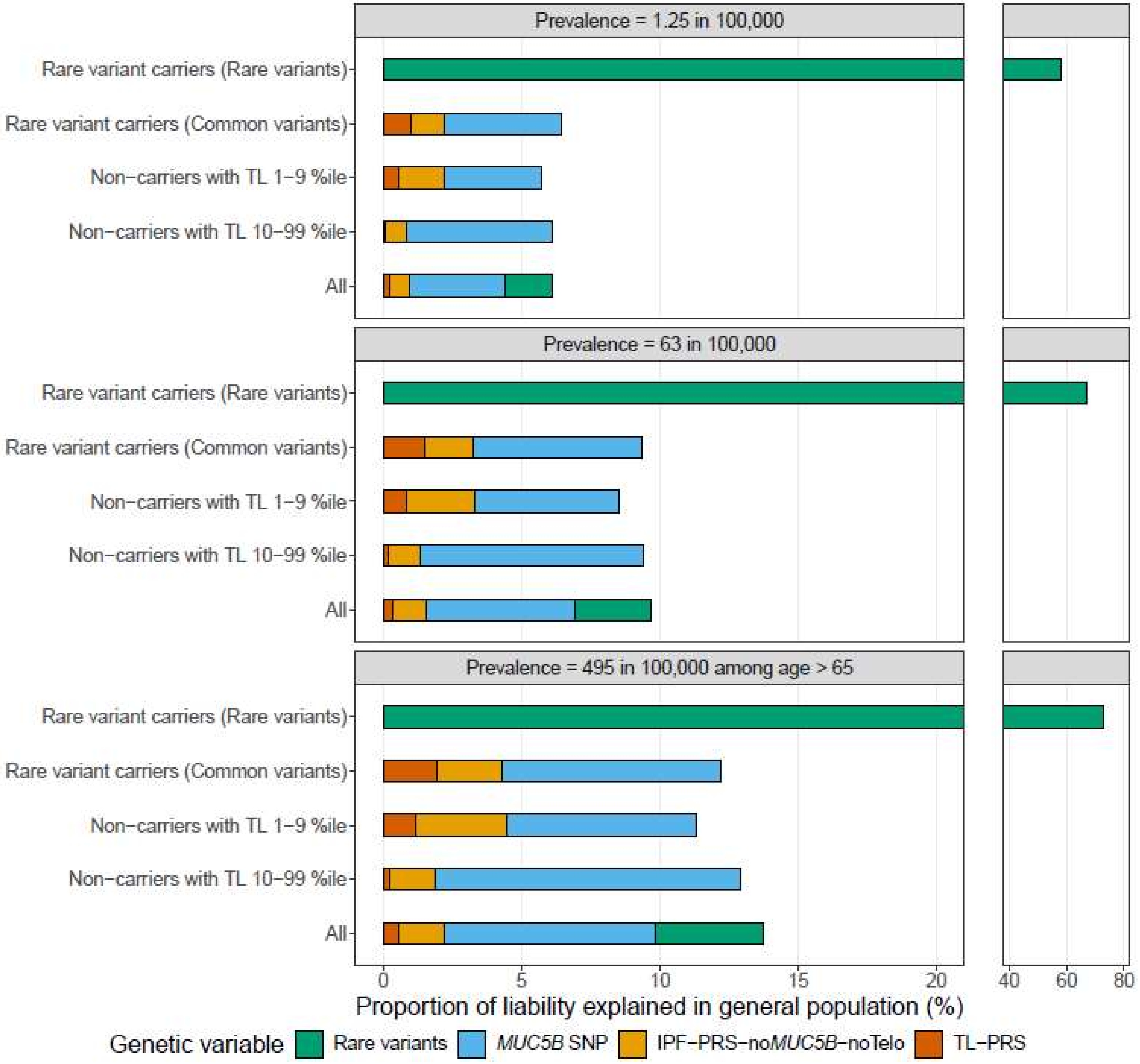


**Supplemental Figure 13**. **Proportion of liability of IPF explained by genetic variables in general population for non-European ancestry individuals.** Data shown using Columbia cohort. Liability explained across range of true prevalences of IPF in the general population as well as in individuals over age 65.

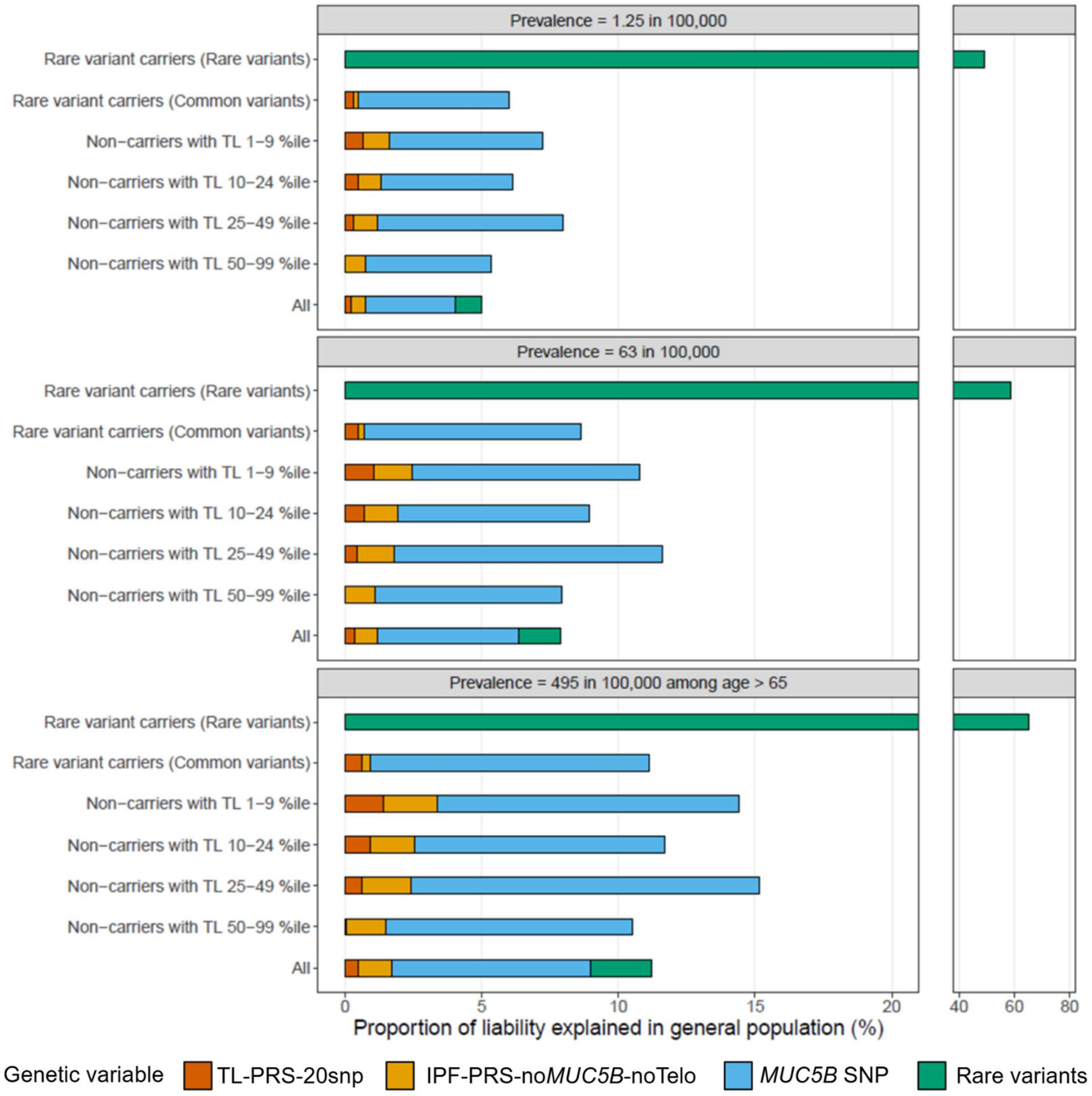


**Supplemental Figure 14**. **Proportion of liability of IPF explained by genetic variables using alternative 20-SNP telomere length polygenic score in the general population.** Data shown using Columbia cohort. Liability explained across range of true prevalences of IPF in the general population as well as in individuals over age 65. Telomere length polygenic score computed using alternative telomere length GWAS with 20 genome-wide significant SNPs from Li et al.

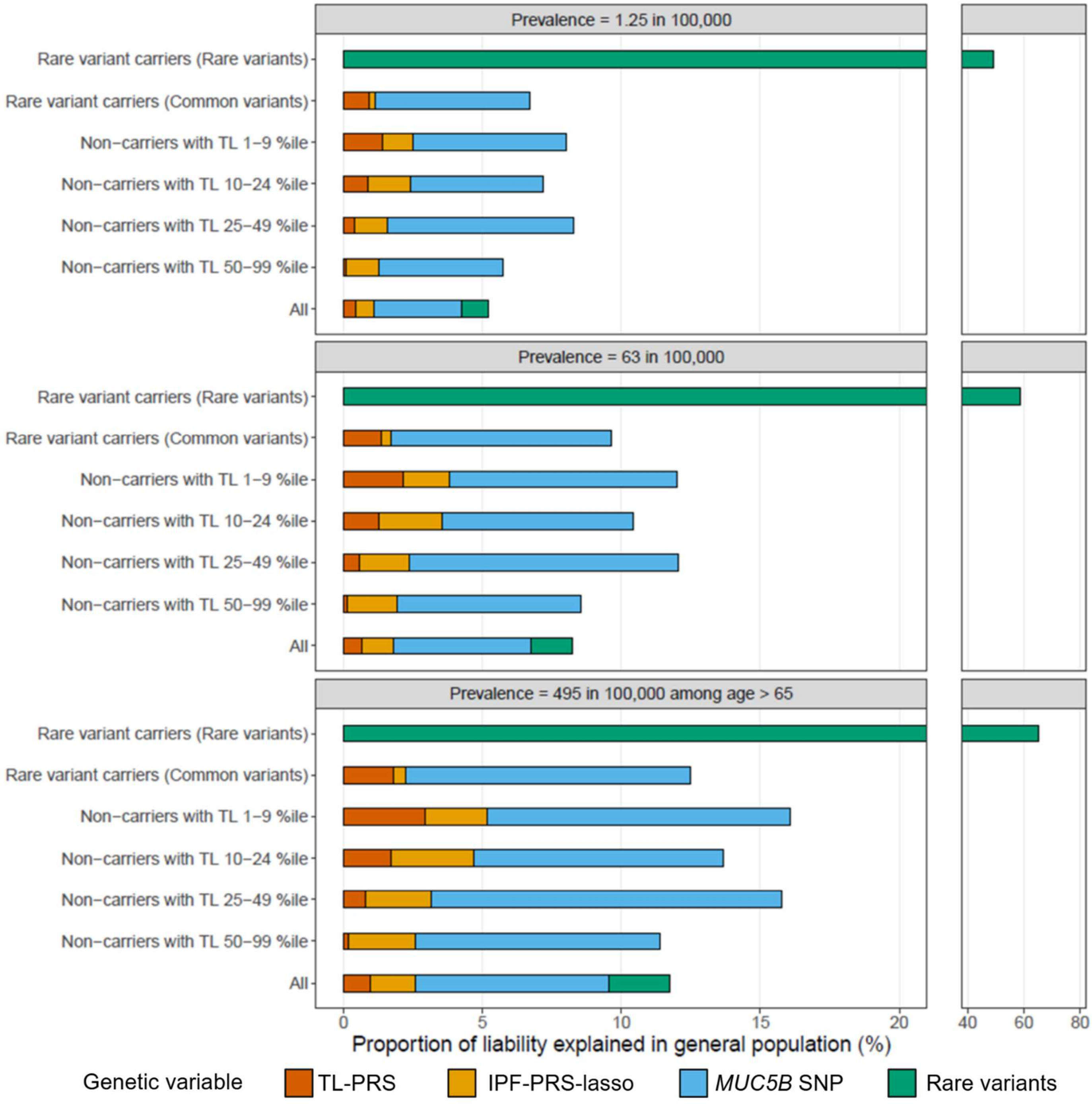


**Supplemental Figure 15**. **Proportion of liability of IPF explained by genetic variables using alternative lasso regression IPF polygenic score in the general population.** Data shown using Columbia cohort. Liability explained across range of true prevalences of IPF in the general population as well as in individuals over age 65. IPF polygenic score computed using alternative approach using lasso regression from Moll et al[20].

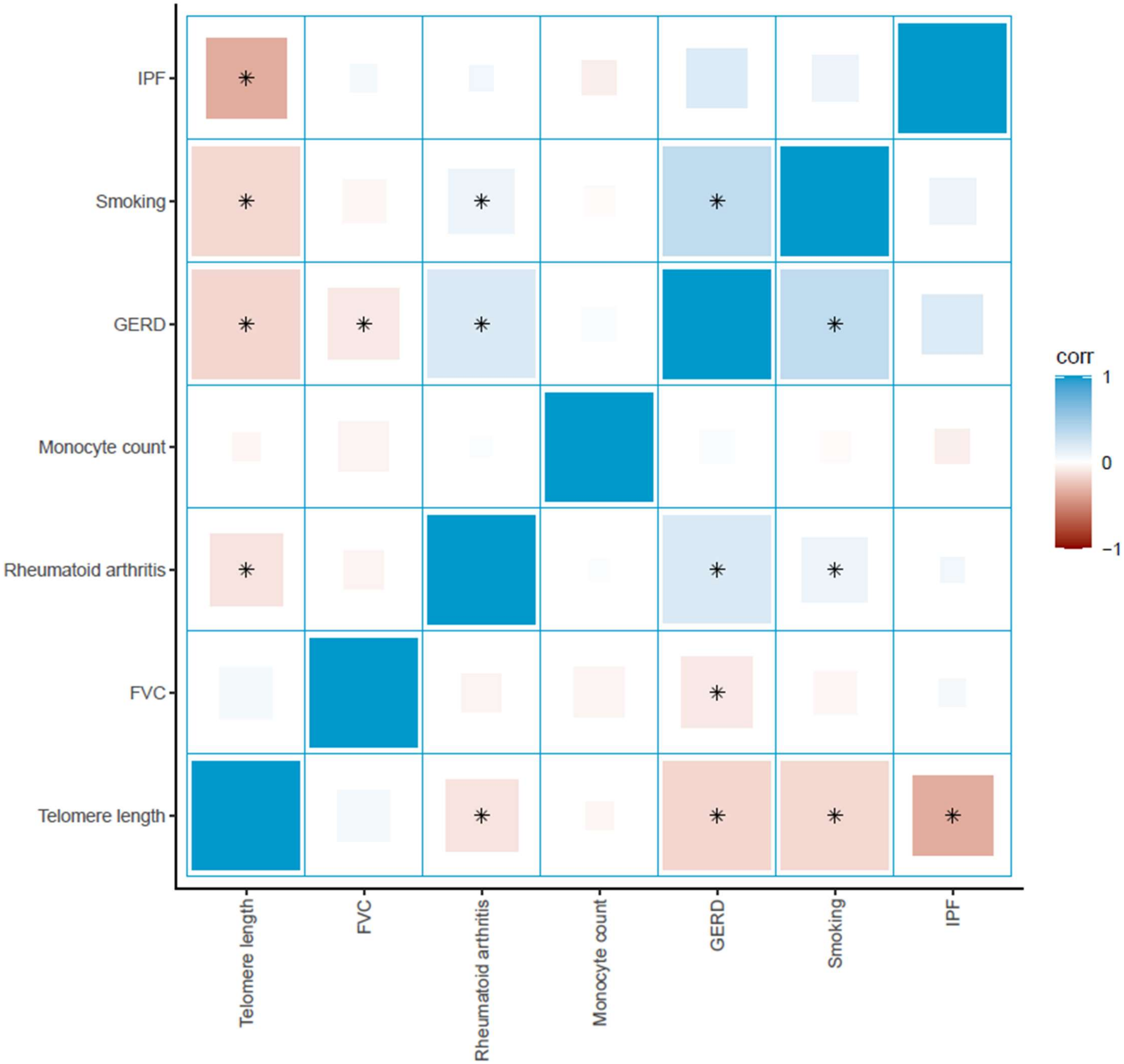


**Supplemental Figure 16. Pair-wise genetic correlation amongst IPF and epidemiologically associated traits.** Genetic correlation estimated from existing GWAS summary statistics of IPF[18], telomere length[19], and other epidemiologically associated traits[22-26] using cross-trait linkage-disequilibrium score regression. Size of box indicates significance of correlation. Shade of box color indicates genetic correlation coefficient (R). Starred boxes indicate pair-wise comparisons with significance that exceeds Bonferroni-corrected threshold of 0.05/21 independent tests. Only telomere length had significant genetic correlation with IPF amongst all other tested traits ($r=-0.35$, $p=3.6\times10^{-6}$); decrease in genetically predicted telomere length correlated with increased genetic risk of IPF.

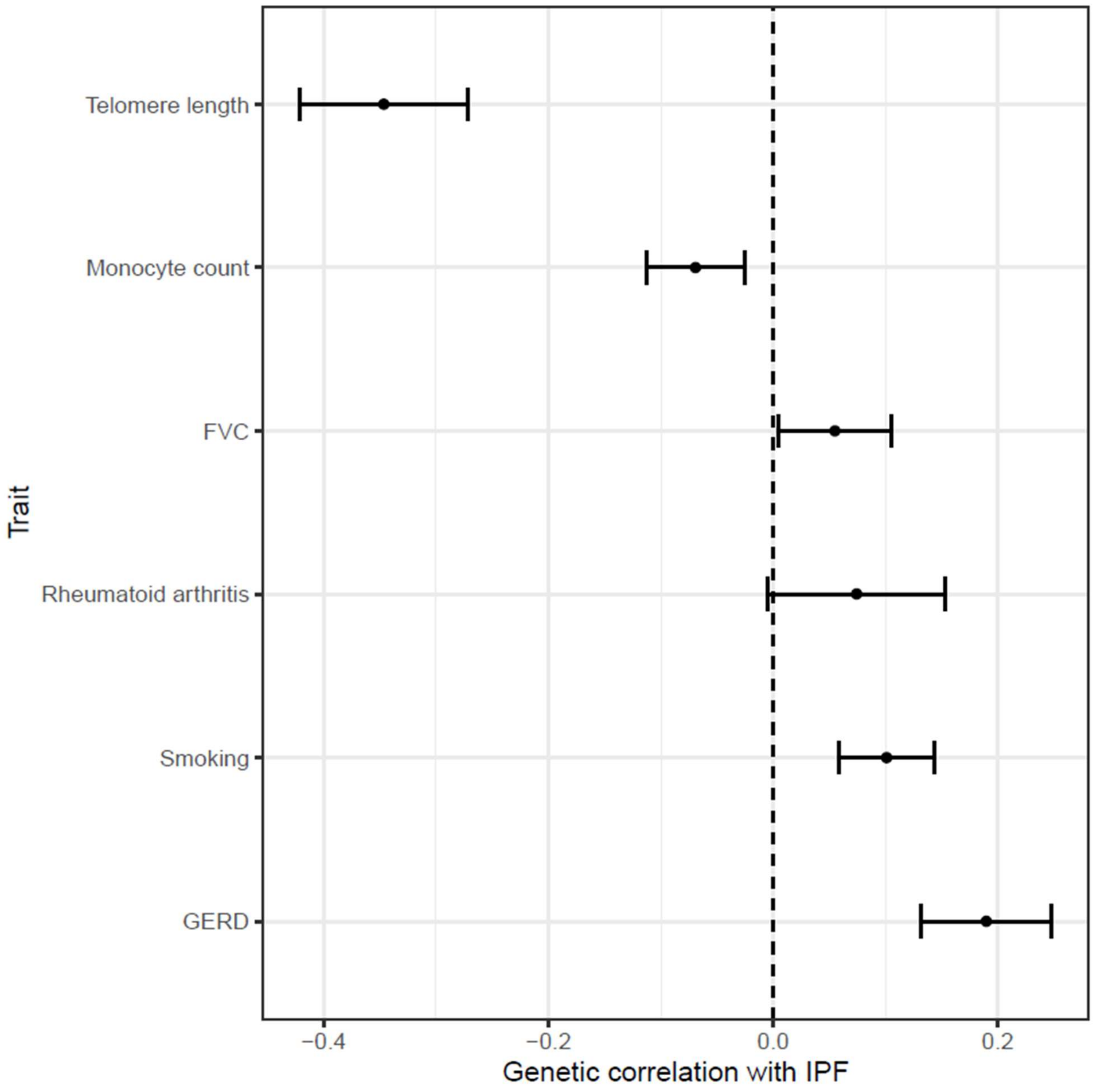


**Supplemental Figure 17**. **Genetic correlation between IPF and epidemiologically associated traits.** Correlation calculated using summary statistics of GWAS studies for each trait of only European individuals. Points represent genetic correlation coefficient (R) with 95% confidence intervals. Genetically shorter telomere length is the most correlated with increased genetic risk of IPF compared to other traits.

**Supplemental Table S1.** Genome-wide significant SNPs associated with IPF

| rsid | gene | chr | hg19pos | hg38pos | EA | EAF | beta |
|---|---|---|---|---|---|---|---|
| rs78238620 | KIF15 | 3 | 44902386 | 44860894 | A | 0.053 | 0.457425 |
| rs12696304 | LRRC34/TERC | 3 | 169481271 | 169763483 | G | 0.279 | 0.270027 |
| rs2013701 | FAM13A | 4 | 89885086 | 88963935 | T | 0.487 | -0.24846 |
| rs7725218 | TERT | 5 | 1282414 | 1282299 | A | 0.325 | -0.3285 |
| rs2076295 | DSP | 6 | 7563232 | 7562999 | G | 0.469 | 0.378436 |
| rs12699415 | MAD1L1 | 7 | 1909479 | 1869843 | A | 0.42 | 0.24686 |
| rs2897075 | 7q22.1 | 7 | 99630342 | 100032719 | T | 0.391 | 0.262364 |
| rs28513081 | DEPTOR | 8 | 120934126 | 119921886 | G | 0.428 | -0.19845 |
| rs537322302 | HECTD2 | 10 | 93271016 | 91511259 | G | 0.003 | 2.056685 |
| rs35705950 | MUC5B | 11 | 1241221 | 1219991 | T | 0.149 | 1.576915 |
| rs9577395 | ATP11A | 13 | 113534984 | 112880670 | G | 0.207 | -0.26136 |
| rs59424629 | IVD | 15 | 40720542 | 40428343 | G | 0.461 | -0.26136 |
| rs62023891 | AKAP13 | 15 | 86097216 | 85553985 | A | 0.3 | 0.239017 |
| rs2077551 | MAPT | 17 | 44214888 | 46137522 | C | 0.186 | -0.34249 |
| rs12610495 | DPP9 | 19 | 4717672 | 4717660 | G | 0.305 | 0.270027 |
| rs41308092 | RTEL1 | 20 | 62324391 | 63693038 | A | 0.021 | 0.751416 |

Betas for each SNP from published IPF GWAS[18]. EA, effect allele; EAF, effect allele frequency

**Supplemental Table S2.** Genome-wide significant autosomal SNPs associated with telomere length

| rsid | gene | chr | hg19pos | hg38pos | EA | EAF | beta |
|---|---|---|---|---|---|---|---|
| rs187540244 | EXOSC10 | 1 | 11224327 | 11164270 | G | 0.995418 | 0.091 |
| rs66731853 | CDA | 1 | 20916238 | 20589745 | G | 0.682696 | 0.018 |
| rs17185038 | RPA2 | 1 | 28219658 | 27893147 | C | 0.93539 | 0.026 |
| rs6669563 | SPOCD1 | 1 | 32279629 | 31814028 | G | 0.562232 | -0.018 |
| rs3768321 | PABPC4 | 1 | 40035928 | 39570256 | G | 0.810305 | 0.016 |
| 1:41236837_CT_C | NFYC | 1 | 41236837 | 40771165 | CT | 0.774903 | -0.015 |
| rs41269079 | BEST4 | 1 | 45252015 | 44786343 | T | 0.811009 | -0.015 |
| rs139795227 | RPAP2 | 1 | 92842367 | 92376810 | A | 0.985979 | -0.06 |
| rs4498805 | SLC16A4 | 1 | 110910397 | 110367775 | G | 0.453368 | -0.015 |
| rs3838300 | MAGI3 | 1 | 114442355 | 113899742 | C | 0.819725 | 0.033 |
| rs11579626 | CHD1L | 1 | 146741960 | 147270300 | A | 0.915118 | -0.027 |
| rs61818036 | PSMB4 | 1 | 151364199 | 151391723 | G | 0.176391 | 0.019 |
| rs932002 | PARP1 | 1 | 226577306 | 226389605 | C | 0.849157 | 0.04 |
| rs9752694 | SMC6 | 2 | 17874177 | 17692910 | C | 0.571283 | 0.014 |
| rs56178008 | TRMT61B | 2 | 29098543 | 28875677 | T | 0.562503 | -0.014 |
| rs12615793 | ACYP2 | 2 | 54475914 | 54248777 | G | 0.859605 | -0.045 |
| rs12613375 | LINC01122 | 2 | 58984109 | 58756974 | C | 0.862288 | -0.018 |
| rs775145631 | UNC80 | 2 | 210667432 | 209802708 | TC | 0.409029 | 0.027 |
| rs35671754 | ATIC | 2 | 216220870 | 215356147 | G | 0.704253 | -0.012 |
| rs869785 | THRB | 3 | 24347800 | 24306309 | T | 0.327529 | 0.015 |
| rs575032615 | SMARCC1 | 3 | 47638657 | 47597167 | A | 0.984904 | -0.061 |
| 3:49959570_CA_C | MST1R | 3 | 49959570 | 49922137 | CA | 0.483013 | 0.016 |
| rs78491606 | SHQ1 | 3 | 72891547 | 72842396 | A | 0.981567 | 0.076 |

| | | | | | | | |
|---|---|---|---|---|---|---|---|
| rs13062095 | SENP7 | 3 | 101267385 | 101548541 | T | 0.672157 | -0.014 |
| rs6776756 | GATA2 | 3 | 128215821 | 128496978 | G | 0.402438 | 0.017 |
| rs2811491 | GATA2 | 3 | 128353950 | 128635107 | C | 0.39223 | -0.015 |
| 3:138398778_TA_T | PIK3CB | 3 | 138398778 | 138679936 | TA | 0.44132 | -0.015 |
| rs201009932 | SMC4 | 3 | 160086718 | 160368930 | T | 0.991068 | 0.077 |
| rs41272947 | SMC4 | 3 | 160119525 | 160401737 | G | 0.458021 | 0.017 |
| rs2293607 | TERC | 3 | 169482335 | 169764547 | T | 0.757553 | 0.094 |
| rs146546514 | TERC | 3 | 169553070 | 169835282 | C | 0.983095 | -0.08 |
| rs753936006 | POLN | 4 | 2191750 | 2190023 | CAAAAA | 0.886351 | 0.025 |
| rs871134 | CCDC96 | 4 | 7044380 | 7042653 | C | 0.430968 | 0.018 |
| rs13129697 | SLC2A9 | 4 | 9926967 | 9925343 | T | 0.720832 | -0.017 |
| rs4695407 | OCIAD1 | 4 | 48843372 | 48841355 | A | 0.492157 | -0.014 |
| rs35500378 | EXOSC9 | 4 | 122729413 | 121808258 | CACTT | 0.610542 | 0.014 |
| rs4435700 | NAF1 | 4 | 164020174 | 163099022 | C | 0.235067 | -0.054 |
| rs113580095 | NAF1 | 4 | 164065720 | 163144568 | A | 0.998088 | 0.37 |
| rs72631678 | NAF1 | 4 | 164102715 | 163181563 | G | 0.587926 | -0.034 |
| rs9990898 | NAF1 | 4 | 164149124 | 163227972 | T | 0.857338 | -0.03 |
| rs112951499 | LOC105374602 | 5 | 50697 | 50593 | A | 0.936584 | -0.034 |
| rs138895564 | TERT | 5 | 1272074 | 1271959 | C | 0.990903 | -0.18 |
| rs575928023 | TERT | 5 | 1285213 | 1285098 | C | 0.989953 | -0.091 |
| rs7705526 | TERT | 5 | 1285974 | 1285859 | C | 0.673422 | -0.078 |
| rs112290073 | TERT | 5 | 1286032 | 1285917 | G | 0.988482 | -0.14 |
| rs2853677 | TERT | 5 | 1287194 | 1287079 | G | 0.424245 | 0.055 |
| rs115451758 | TERT | 5 | 1289277 | 1289162 | G | 0.989626 | 0.084 |
| rs34094720 | TERT | 5 | 1293767 | 1293652 | G | 0.994776 | -0.16 |
| rs61748181 | TERT | 5 | 1294166 | 1294051 | C | 0.971072 | 0.059 |
| rs33987166 | TERT | 5 | 1296758 | 1296643 | T | 0.975189 | -0.098 |
| 5:1303867_CT_C | TERT | 5 | 1303867 | 1303752 | CT | 0.994812 | -0.14 |
| rs79717857 | TERT | 5 | 1319997 | 1319882 | C | 0.973921 | -0.051 |
| rs115251750 | TERT | 5 | 1349864 | 1349749 | G | 0.964935 | -0.036 |
| rs113206288 | TERT | 5 | 1611683 | 1611568 | A | 0.85294 | -0.023 |
| 5:78951569_GT_G | TENT2 (PAPD4) | 5 | 78951569 | 79655746 | GT | 0.898518 | 0.025 |
| rs72801474 | HSPA4 | 5 | 132444128 | 133108436 | G | 0.911707 | 0.021 |
| rs34255404 | UBE2D2 | 5 | 138935580 | 139555995 | G | 0.943277 | -0.037 |
| rs80324517 | LOC285766 | 6 | 204031 | 204031 | G | 0.951741 | -0.04 |
| rs55965437 | PRRC2A | 6 | 31797769 | 31829992 | A | 0.640531 | 0.028 |
| rs154979 | HLA-DMB | 6 | 32890939 | 32923162 | C | 0.971166 | -0.044 |
| rs9398196 | CCDC162P | 6 | 109601554 | 109280351 | A | 0.47995 | 0.014 |
| rs117247304 | LOC102723672 | 7 | 69010 | 69010 | G | 0.975555 | 0.064 |
| rs13230646 | STK31 | 7 | 23930316 | 23890697 | T | 0.751055 | 0.017 |
| rs11769630 | IKZF1 | 7 | 50257703 | 50218107 | T | 0.927773 | 0.026 |
| rs2538745 | UPK3B | 7 | 76310784 | 76681467 | T | 0.397159 | 0.013 |
| rs2056726 | STAG3 | 7 | 99780283 | 100182660 | G | 0.785624 | 0.023 |
| rs609953 | RNU6-2 | 7 | 123422444 | 123782390 | T | 0.617692 | -0.013 |
| rs7790856 | POT1 | 7 | 124459852 | 124819798 | C | 0.710861 | 0.044 |

| | | | | | | | |
|---|---|---|---|---|---|---|---|
| rs117811540 | POT1 | 7 | 124569916 | 124929862 | G | 0.991965 | -0.142 |
| rs36101328 | POT1 | 7 | 124601146 | 124961107 | C | 0.514892 | 0.031 |
| rs4731541 | TNPO3 | 7 | 128678236 | 129038182 | C | 0.375099 | 0.021 |
| rs11556924 | ZC3HC1 | 7 | 129663496 | 130023656 | C | 0.624146 | -0.013 |
| rs138061125 | VIPR2 | 7 | 159079192 | 159286503 | G | 0.969815 | -0.043 |
| rs1985369 | VIPR2 | 7 | 159119220 | 159326530 | A | 0.131822 | 0.031 |
| rs2306646 | XPO7 | 8 | 21846586 | 21989075 | G | 0.440525 | 0.021 |
| rs762679 | MCM4 | 8 | 48885436 | 47972876 | T | 0.143499 | -0.031 |
| rs564224004 | TGS1 | 8 | 56667353 | 55754800 | C | 0.1149 | 0.031 |
| rs7012816 | PRDM14 | 8 | 70964743 | 70052508 | G | 0.870003 | -0.018 |
| rs10112752 | TERF1 | 8 | 73958718 | 73046483 | G | 0.569631 | 0.029 |
| rs540491189 | TERF1 | 8 | 74150379 | 73238144 | G | 0.997823 | -0.191 |
| rs1023767 | VIRMA | 8 | 95530969 | 94518741 | G | 0.762405 | 0.018 |
| rs4742448 | DMRT1 | 9 | 826585 | 826585 | C | 0.530842 | -0.015 |
| rs11557154 | DCAF12 | 9 | 34107505 | 34107507 | C | 0.86997 | 0.034 |
| rs4743037 | ZNF462 | 9 | 109639970 | 106877689 | C | 0.769126 | -0.015 |
| rs762222726 | ASB13 | 10 | 5816070 | 5774107 | CAAACAT | 0.399185 | 0.018 |
| rs12572897 | NOC3L | 10 | 96114835 | 94355078 | G | 0.869908 | 0.032 |
| 10:101274251_CT_C | NKX2-3 | 10 | 101274251 | 99514494 | CT | 0.619614 | 0.022 |
| rs11190184 | NKX2-3 | 10 | 101368199 | 99608442 | G | 0.71576 | 0.017 |
| rs4919611 | PPRC1 | 10 | 103894939 | 102135182 | C | 0.112898 | 0.026 |
| rs10748858 | STN1 (OBFC1) | 10 | 105639514 | 103879756 | T | 0.595083 | -0.039 |
| rs9419958 | STN1 (OBFC1) | 10 | 105675946 | 103916188 | T | 0.13861 | 0.081 |
| rs182641927 | STN1 (OBFC1) | 10 | 105805039 | 104045281 | C | 0.995952 | -0.196 |
| rs939916 | ODF3 | 11 | 202253 | 202253 | G | 0.330033 | -0.024 |
| rs10840270 | WEE1 | 11 | 9629553 | 9608006 | C | 0.344316 | -0.014 |
| rs2293579 | PSMC3 | 11 | 47440758 | 47419207 | G | 0.613726 | 0.013 |
| rs141379009 | ATM | 11 | 108149207 | 108278480 | T | 0.974488 | 0.092 |
| rs611646 | ATM | 11 | 108177097 | 108306370 | T | 0.591315 | 0.037 |
| rs6590343 | FLI1 | 11 | 128500215 | 128630320 | A | 0.483615 | -0.012 |
| rs10845387 | LINC01252 | 12 | 11757743 | 11604809 | G | 0.647334 | 0.014 |
| rs12369950 | LINC00477 | 12 | 24762109 | 24609175 | T | 0.859325 | 0.018 |
| rs79977579 | SMUG1 | 12 | 54694560 | 54300776 | C | 0.90445 | -0.028 |
| rs1907702 | KITLG | 12 | 88955469 | 88561692 | G | 0.233229 | -0.015 |
| rs10774625 | SH2B3 | 12 | 111910219 | 111472415 | A | 0.476428 | -0.016 |
| rs76666449 | SRSF9 | 12 | 120904895 | 120467092 | T | 0.899375 | -0.03 |
| rs4758644 | ZCCHC8 | 12 | 122943915 | 122459368 | A | 0.269222 | 0.017 |
| rs1727302 | MPHOSPH9 | 12 | 123632930 | 123148383 | G | 0.259019 | -0.02 |
| rs79228077 | MUC8 | 12 | 133046343 | 132469757 | G | 0.368978 | 0.015 |
| rs1332941 | KBTBD6 | 13 | 41695100 | 41120964 | A | 0.179534 | -0.026 |
| rs35017269 | DIS3 | 13 | 73340177 | 72766039 | G | 0.984244 | -0.067 |
| rs3093888 | TEP1 | 14 | 20812951 | 20344792 | G | 0.94869 | 0.029 |
| rs73581419 | RAB2B | 14 | 21941148 | 21472989 | C | 0.893395 | -0.023 |
| rs12884911 | PPP1R36 | 14 | 65027871 | 64561153 | C | 0.49667 | 0.013 |
| rs762810 | MAX | 14 | 65544367 | 65077649 | C | 0.647514 | 0.02 |
| rs137901416 | DCAF4 | 14 | 73418095 | 72951387 | G | 0.899689 | -0.046 |

| | | | | | | | |
|---|---|---|---|---|---|---|---|
| rs1007934 | DCAF4 | 14 | 73463479 | 72996771 | G | 0.594615 | 0.023 |
| 14:91970514_GA_G | PPP4R3A | 14 | 91970514 | 91504170 | GA | 0.544128 | -0.019 |
| rs1957937 | TCL1A | 14 | 96181360 | 95715023 | A | 0.83982 | -0.021 |
| rs17677991 | MGA | 15 | 42032383 | 41740185 | C | 0.657877 | -0.022 |
| rs181647350 | ATP8B4 | 15 | 50379219 | 50087022 | T | 0.761155 | 0.034 |
| rs1980240 | TEX9 | 15 | 56774018 | 56481820 | A | 0.594757 | -0.013 |
| rs5742915 | PML | 15 | 74336633 | 74044292 | T | 0.554171 | -0.019 |
| rs80116508 | SLX4 | 16 | 3650970 | 3600969 | G | 0.937644 | 0.035 |
| rs11646283 | USP7 | 16 | 9073060 | 8979203 | T | 0.589297 | -0.015 |
| rs182059586 | PARN | 16 | 14652220 | 14558363 | T | 0.97489 | 0.057 |
| rs450962 | EIF3CL | 16 | 28413517 | 28402196 | A | 0.716221 | -0.014 |
| rs8053839 | LONP2 | 16 | 48390512 | 48356601 | G | 0.458038 | 0.014 |
| rs12447324 | PAPD5 | 16 | 50089038 | 50055127 | C | 0.803162 | -0.017 |
| rs76219171 | PAPD5 | 16 | 50188929 | 50155018 | G | 0.941567 | -0.036 |
| rs28711261 | ACD | 16 | 67617186 | 67583283 | A | 0.867449 | -0.017 |
| rs139438549 | ACD | 16 | 67692863 | 67658960 | T | 0.998884 | -0.256 |
| rs142507451 | ACD | 16 | 67694044 | 67660141 | C | 0.997058 | 0.142 |
| rs528301822 | TERF2 | 16 | 69403012 | 69369109 | A | 0.710663 | -0.024 |
| rs62053340 | EXOSC6 | 16 | 69987764 | 69953861 | C | 0.617037 | 0.021 |
| rs529549411 | EXOSC6 | 16 | 70390510 | 70356607 | C | 0.994309 | 0.088 |
| rs34003787 | ZFHX3 | 16 | 73071381 | 73037482 | C | 0.912293 | 0.024 |
| rs183553155 | RFWD3 | 16 | 74491239 | 74457341 | G | 0.989437 | -0.071 |
| rs11866592 | RFWD3 | 16 | 74654396 | 74620498 | G | 0.85802 | -0.035 |
| rs7193541 | RFWD3 | 16 | 74664743 | 74630845 | T | 0.583445 | 0.021 |
| rs2303262 | MPHOSPH6 | 16 | 82203758 | 82170153 | C | 0.222887 | 0.047 |
| rs62046862 | BANP | 16 | 88073029 | 88039423 | C | 0.479275 | -0.024 |
| rs35216338 | BANP | 16 | 88103010 | 88069404 | C | 0.960393 | -0.045 |
| rs9923119 | PRDM7 | 16 | 90153815 | 90087407 | T | 0.7498 | -0.018 |
| rs7218033 | RPA1 | 17 | 1694247 | 1790953 | C | 0.746597 | 0.023 |
| rs5030755 | RPA1 | 17 | 1782952 | 1879658 | A | 0.886658 | -0.03 |
| rs4724 | CTC1 | 17 | 7760397 | 7857079 | G | 0.883402 | 0.055 |
| rs75664430 | CTC1 | 17 | 8064779 | 8161461 | C | 0.751972 | 0.024 |
| rs111527438 | ADAP2 | 17 | 29252703 | 30925685 | T | 0.648749 | -0.013 |
| rs62079650 | BRCA1 | 17 | 41401261 | 43323893 | A | 0.265737 | 0.023 |
| rs12941945 | BRCA1 | 17 | 41448228 | 43370860 | A | 0.832437 | 0.026 |
| rs34405642 | NOL11 | 17 | 65741012 | 67744910 | G | 0.414632 | -0.013 |
| rs2069536 | TEN1 | 17 | 74001106 | 76005025 | A | 0.249795 | 0.015 |
| rs1143697 | TK1 | 17 | 76178748 | 78182667 | T | 0.536558 | -0.014 |
| rs144204502 | TK1 | 17 | 76183233 | 78187152 | C | 0.987438 | 0.101 |
| rs9952504 | TYMS | 18 | 657458 | 657458 | A | 0.958066 | 0.053 |
| rs3891167 | TYMS | 18 | 658423 | 658423 | A | 0.746565 | 0.043 |
| rs111811424 | TYMS | 18 | 674579 | 674579 | C | 0.913521 | -0.039 |
| rs2741181 | TYMS | 18 | 689437 | 689437 | C | 0.912964 | 0.048 |
| rs116863223 | TYMS | 18 | 709396 | 709396 | G | 0.988237 | 0.082 |
| rs78694226 | TYMS | 18 | 710980 | 710980 | G | 0.992114 | 0.066 |
| rs79824385 | LINC01478 | 18 | 41982379 | 44402414 | T | 0.870276 | -0.032 |

| | | | | | | | |
|---|---|---|---|---|---|---|---|
| rs8088824 | LINC01478 | 18 | 42151261 | 44571296 | C | 0.236812 | -0.026 |
| rs2276182 | POLI | 18 | 51798047 | 54271677 | C | 0.596773 | -0.023 |
| rs6565924 | ZNF236 | 18 | 74691225 | 76979269 | A | 0.638554 | -0.013 |
| rs1879100 | PARD6G | 18 | 77985740 | 80227857 | C | 0.137732 | 0.019 |
| rs80337039 | MAP2K2 | 19 | 4105089 | 4105091 | G | 0.993522 | -0.081 |
| rs35601737 | TRMT1 | 19 | 13220703 | 13109889 | C | 0.704101 | 0.014 |
| rs8105767 | ZNF208 | 19 | 22215441 | 22032639 | A | 0.70533 | -0.033 |
| rs4530278 | CEBPA | 19 | 33752994 | 33262088 | G | 0.40185 | -0.014 |
| rs429358 | APOE | 19 | 45411941 | 44908684 | T | 0.846031 | -0.017 |
| rs11084431 | ZSCAN5B | 19 | 56708667 | 56197298 | G | 0.395562 | 0.012 |
| rs8102497 | PEG3 | 19 | 57370055 | 56858687 | G | 0.568172 | 0.015 |
| rs1291143 | SAMHD1 | 20 | 35525640 | 36897237 | A | 0.150974 | -0.049 |
| rs112802859 | SAMHD1 | 20 | 35549787 | 36921384 | C | 0.90456 | -0.028 |
| rs6030416 | SAMHD1 | 20 | 35584624 | 36956221 | T | 0.135696 | 0.034 |
| rs544699357 | SRSF6 | 20 | 41997057 | 43368417 | C | 0.998007 | -0.169 |
| rs577449057 | RTEL1 | 20 | 62236709 | 63605356 | A | 0.994723 | 0.154 |
| rs2259797 | RTEL1 | 20 | 62272248 | 63640895 | T | 0.907332 | 0.081 |
| rs41308088 | RTEL1 | 20 | 62293118 | 63661765 | C | 0.918776 | -0.048 |
| rs187013287 | RTEL1 | 20 | 62298374 | 63667021 | A | 0.99742 | 0.283 |
| rs8114049 | RTEL1 | 20 | 62310806 | 63679453 | C | 0.330913 | 0.042 |
| rs35640778 | RTEL1 | 20 | 62321128 | 63689775 | G | 0.979243 | 0.209 |
| 20:62321690_GAGA_G | RTEL1 | 20 | 62321690 | 63690337 | GAGA | 0.995641 | -0.118 |
| rs115610405 | RTEL1 | 20 | 62325833 | 63694480 | C | 0.981137 | 0.11 |
| rs55765053 | RTEL1 | 20 | 62328480 | 63697127 | C | 0.933938 | -0.022 |
| rs3761121 | RTEL1 | 20 | 62342695 | 63711343 | T | 0.878032 | -0.02 |
| rs187577818 | RTEL1 | 20 | 62358869 | 63727517 | A | 0.997306 | -0.298 |
| rs111527478 | RTEL1 | 20 | 62678100 | 64046747 | G | 0.900225 | -0.022 |
| rs28502153 | GAB4 | 22 | 17469049 | 16988159 | C | 0.622042 | 0.022 |
| rs5845706 | SMC1B | 22 | 45779013 | 45383144 | C | 0.643742 | -0.015 |
| rs131796 | TYMP | 22 | 50971639 | 50533219 | G | 0.235626 | 0.024 |

Betas for each SNP from published telomere length GWAS[19]. EA, effect allele; EAF, effect allele frequency

**Supplemental Table S3.** Overlapping genetic loci in GWAS studies of IPF and telomere length

| IPF GWAS SNP | Telomere length GWAS SNP | Gene Loci | Linkage Disequilibrium $R^2$ (Europeans) | IPF GWAS p-value | TL GWAS p-value |
|---|---|---|---|---|---|
| rs7725218 | rs7705526 | *TERT* | 0.71 | $1.5 \times 10^{-20}$ | $9.1 \times 10^{-272}$ |
| rs12696304 | rs2293607 | *TERC* | 0.88 | $7.1 \times 10^{-13}$ | $<1 \times 10^{-314}$ |
| rs41308092 | rs115610405 | *RTEL1* | 0.76 | $7.7 \times 10^{-10}$ | $4.6 \times 10^{-51}$ |

Published genome-wide significant loci for GWAS of IPF[18] and of telomere length[19] in linkage disequilibrium with respective published p-values

**Supplemental Table S4.** Descriptions of polygenic scores

| Polygenic score name | Trait | *MUC5B* rs35705950 | Telomere-associated SNPs | No. SNPs | GWAS for generating score | Used for analysis |
|---|---|---|---|---|---|---|
| IPF-PRS-*MUC5B* | IPF | Included | Included | 16 | Allen et al.[18] | Sensitivity analysis |
| IPF-PRS-no*MUC5B*-withTelo | IPF | Excluded | Included | 15 | Allen et al.[18] | Sensitivity analysis |
| **IPF-PRS-no*MUC5B*-noTelo** | **IPF** | **Excluded** | **Excluded** | **12** | **Allen et al.[18]** | **Primary analysis** |
| IPF-PRS-lasso | IPF | Excluded | Included | 60,608 | Allen et al.[18] | Sensitivity analysis |
| **TL-PRS** | **TL** | **-** | **Included** | **190** | **Codd et al.[19]** | **Primary analysis** |
| TL-PRS-noTelo | TL | - | Excluded | 187 | Codd et al.[19] | Sensitivity analysis |
| TL-PRS-20snp | TL | - | Included | 20 | Li et al.[21] | Sensitivity analysis |

**Supplemental Table S5.** Phenotypes of controls in Columbia discovery cohort.

| Control Phenotypes | No. Samples |
|---|---|
| Kidney and urologic disease | 1772 (61%) |
| Amyotrophic lateral sclerosis | 701 (24%) |
| COVID-19 | 428 (15%) |
| Other | 4 (<1%) |
| **Total** | **2905 (100%)** |

**Supplemental Table S6.** Age, sex, and ancestry of cohort participants.

| Characteristic | Columbia | | TOPMed | | UKBB | |
|---|---|---|---|---|---|---|
| | IPF (n=777) | Controls (n=2905) | IPF (n=1148) | Controls (n=5202) | IPF (n=2739) | Controls (n=395331) |
| Age (median, IQR) | 67 (61, 73) | 55 (41, 70) | 66 (60, 72) | 63 (55, 71) | 72 (67, 77)*<br>64 (61, 68)& | 58 (52, 65)& |
| Male sex, No. (%) | 562 (72%) | 1665 (57%) | 796 (69%) | 2504 (48%) | 1674 (61%) | 181704 (46%) |
| Genetic ancestry, No. (%) | | | | | | |
| European | 656 (84%) | 1995 (69%) | 1100 (96%) | 2800 (54%) | 2626 (96%) | 363558 (95%) |
| African | 22 (3%) | 331 (11%) | 9 (<1%) | 1025 (20%) | 35 (1%) | 7374 (2%) |
| American Admixed | 74 (10%) | 342 (12%) | 15 (1%) | 750 (14%) | 3 (<1%) | 459 (<1%) |
| East Asian | 9 (1%) | 106 (4%) | 13 (1%) | 565 (11%) | 7 (<1%) | 2467 (<1%) |
| South Asian | 9 (1%) | 60 (2%) | 4 (<1%) | 1 (<1%) | 68 (2%) | 9343 (2%) |
| Other | 7 (1%) | 71 (2%) | 7 (<1%) | 61 (1%) | - | - |

Ancestry inferred from genetic data using Peddy. TOPMed, Trans-Omics for Precision Medicine; UKBB, UK Biobank. For IPF cases in UKBB, age is shown as *age at diagnosis for comparison with other IPF cohorts and **age at registration for comparison with UKBB controls.

**Supplemental Table S7.** Genes represented by rare variant carriers in IPF cohorts.

| Gene | Columbia (IPF rare variant carrier n=94) | TOPMed (IPF rare variant carrier n=143) | UKBB (IPF rare variant carrier n=108) |
|---|---|---|---|
| *TERT* | 41 (44%) | 60 (45%) | 16 (15%) |
| *RTEL1* | 12 (13%) | 30 (23%) | 27 (25%) |
| *PARN* | 12 (13%) | 16 (12%) | 23 (21%) |
| *KIF15* | 9 (10%) | 7 (6%) | 22 (20%) |
| *TERC* | 6 (6%) | 10 (8%) | 9 (8%) |
| *SFTPC* | 5 (5%) | 6 (5%) | 0 |
| *NAF1* | 4 (4%) | 3 (2%) | 0 |
| *DKC1* | 2 (2%) | 0 | 0 |
| *TINF2* | 0 | 2 (2%) | 2 (2%) |
| *SFTPA1/2* | 1 (1%) | 4 (3%) | 5 (5%) |
| Two genes* | 2 (2%) | 5 (4%) | 4 (4%) |

*Individuals with two rare variants in Columbia cohort (*TERT/KIF15*, *TINF2/SFTPC*), TOPMed cohort (*RTEL1/KIF15, TERT/PARN, TERT/RTEL1, TERT/SFTPA1, PARN/SFTPA1*), UKBB cohort (*TERT/KIF15, RTEL1/KIF15, RTEL1/KIF15, SFTPA2/KIF15*)

**Supplemental Table S8.** Characteristics of IPF endotypes in all cohorts.

| Characteristic | Rare variant carriers[#] | Non-carriers with short TL <10th percentile | Other non-carriers[$] |
|---|---|---|---|
| *Columbia cohort* | | | |
| Age, median (IQR) | 62 (56, 68) | 68 (63, 73) | 69 (63, 75) |
| Male sex, No. (%) | 65 (69%) | 184 (77%) | 313 (70%) |
| European ancestry, No. (%) | 78 (83%) | 213 (89%) | 365 (82%) |
| Total, No. (%) | 94 (100%) | 238 (100%) | 444 (100%) |
| *TOPMed cohort* | | | |
| Age, median (IQR) | 63 (57, 69) | - | 66 (60, 72) |
| Male sex, No. (%) | 89 (61%) | - | 707(71%) |
| European ancestry, No. (%) | 143 (98%) | - | 957 (96%) |
| Total, No. (%) | 146 (100%) | - | 1002 (100%) |
| *UKBB cohort* | | | |
| Age (diagnosis), median (IQR) | 72 (66, 77) | 71 (67, 76) | 72 (67, 78) |
| Age (registration), median (IQR) | 64 (61, 67) | 64 (60, 68) | 65 (61,68) |
| Male sex, No. (%) | 62 (57%) | 357(70%) | 1255 (59%) |
| European ancestry, No. (%) | 94 (87%) | 496 (97%) | 2036 (96%) |
| Total, No. (%) | 108 (100%) | 512 (100%) | 2119 (100%) |

Ages shown at study registration or enrollment unless otherwise specified. Telomere length percentiles are age-adjusted to nomogram of non-diseased individuals. [#]Carriers of rare damaging variants in IPF-associated genes (*TERT, TERC, PARN, RTEL1, DKC1, TINF2, NAF1, SFTPC, SFTPA1/2, KIF15*). [$]Other non-carriers group in TOPMed cohort represents all non-carriers since telomere length data was not available. TL, telomere length; TOPMed, Trans-Omics for Precision Medicine; UKBB, UK Biobank.

**Supplemental Material References**
